# Circulating Senescence Protein Links Exercise Adaptation to Health Outcomes

**DOI:** 10.64898/2026.02.09.26345899

**Authors:** Nicholas Houstis, Qiulian Zhou, Yuling Chen, Sonja Mittag, Vinita Chowdhury, Chao Wu, Meixi Quan, Azrul Kadir, Giulia Guerra, Sashi Weerawarana, Danielle Szczesniak, Justin Guerra, James Rhee, J. Sawalla Guseh, Haobo Li, Bo Li, Aurel B. Leuchtmann, Jorge Ruas, Ulrik Wisloff, Dorthe Stensvold, Anthony Rosenzweig

## Abstract

Adaptation to physiological stress is fundamental to health but varies widely among individuals. In humans, this heterogeneity is evident in markedly different gains in fitness in response to identical exercise training. The molecular determinants of this variable “trainability” remain poorly understood. Here we identify insulin-like growth factor binding protein-7 (IGFBP7), a senescence-associated secreted protein, as a circulating constraint on exercise adaptation. Plasma proteomics in older adults enrolled in a randomized exercise trial revealed that IGFBP7 levels inversely predicted fitness gains after one year of high-intensity interval training despite similar baseline fitness. In mice, genetic deletion of IGFBP7 markedly amplified training-induced gains in exercise capacity across distinct training protocols, whereas somatic overexpression abolished this advantage. In the UK Biobank, lower IGFBP7 levels were associated with reduced mortality and multiple incident age-related diseases, mirroring the breadth of ties between fitness and healthspan. Together, these findings identify circulating IGFBP7 as a molecular brake on physiological plasticity in response to exercise, linking training responsiveness, aging biology, and health outcomes.

## Main Text

Exercise training improves cardiorespiratory fitness, an adaptive response with far-reaching health benefits and an archetype of phenotypic plasticity. Fitness reflects the integrated performance of multiple organ systems involved in oxygen uptake, transport, and utilization (*1*), and can be quantified by peak oxygen consumption (peak VO□) during exercise. Impaired fitness is a common and debilitating manifestation of disease, often experienced as breathlessness or reduced stamina, whereas improvements in fitness confer broad functional benefits across the lifespan. Athletes seek it for performance, older adults for independence, and patients with chronic illness for quality of life and survival. Consistent with this central role, peak VO□is among the strongest prognostic markers across diverse diseases, and one of the most powerful predictors of human healthspan and lifespan (*2-4*).

The strong ties between fitness, functional capacity, and health highlight the importance of understanding what governs the magnitude of adaptive responses to exercise. Trainability–an individual’s responsiveness to a given training protocol–varies widely in humans. Moreover, this heterogeneous plasticity is conserved across species such as flies and rodents (*5-7*). Despite its prevalence, the molecular basis of trainability remains poorly understood. In particular, it is unclear whether adaptive responses to the repeated physiological stress of exercise are actively constrained, and whether such constraints could be lifted to enhance fitness and health.

Here, we sought to identify circulating proteins that both predict trainability in older adults and causally influence adaptive responses to exercise, leveraging samples from a randomized, controlled trial of exercise training. Using plasma proteomics, we examined how baseline circulating proteins relate to subsequent fitness gains and health outcomes. We found that trainability is actively restrained by IGFBP7, which helps explain individual variation in response to exercise. If safely targetable, modulating the determinants of trainability could boost the functional and health benefits of physical activity to populations with comorbidities, the elderly, and the frail.

## Results

### Trainability

Our study population came from a 5-year randomized trial of exercise training in 1567 Norwegian septuagenarians (mean age 73) conducted from 2012-2017 (*8*). Although the trial showed only a nonsignificant trend toward lower all-cause mortality with high-intensity interval training (HIIT) compared with moderate-intensity exercise and usual care, individual responses to training were strikingly heterogeneous. Importantly, secondary analyses suggested that participants who achieved larger gains in cardiorespiratory fitness also experienced greater clinical benefit (*8, 9*). We focused on participants assigned to the HIIT arm of the trial (Table 1), which produced the largest fitness gains (*8, 10*). These individuals were prescribed two HIIT sessions per week, each consisting of four 4-minute intervals at 90% of peak heart rate, interspersed with 3-minute active recovery periods at 60–70% of peak heart rate (*8*).

**Table 1.**
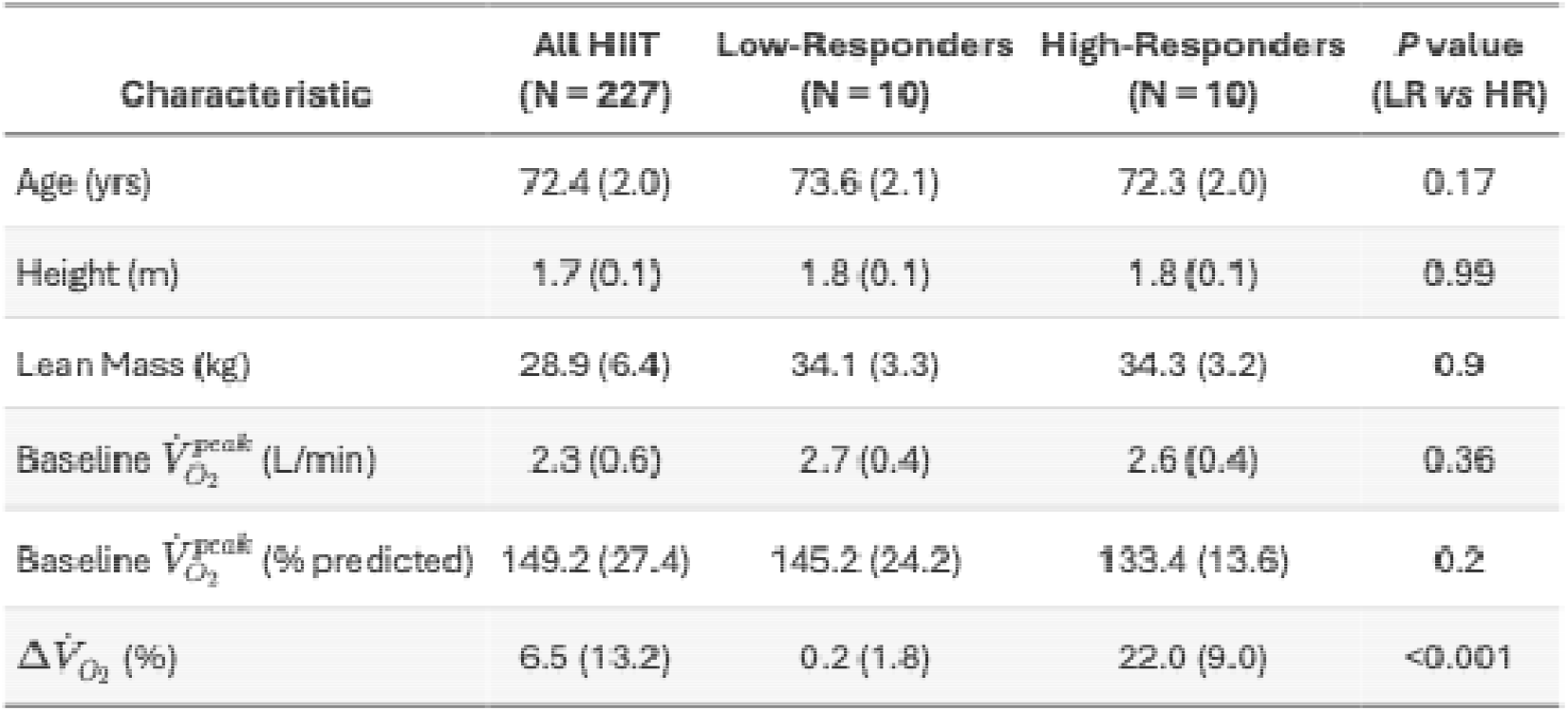
Participant characteristics from the Generation-100 study. Anthropometric and exercise characteristics from the HIIT study arm are reported. Baseline indicates study entry, before exercise training. Training effect (last row) is calculated as the percent increase in VO_2_ over baseline after one year of HIIT. Three groups of participants appear in the table, including the entire HIIT arm as well as two proteomics discovery cohorts of male Low-Responders (LR) and High-Responders (HR). VO_2_ indicates oxygen consumption at peak exercise; HIIT indicates high-intensity interval training.

Although age was narrowly constrained by design (70-77), baseline fitness varied widely, spanning 61% to 240% of predicted peak oxygen consumption (VO_2_). We examined individual adaptive responses to one year of training, including changes in peak VO_2_, resting levels of circulating proteins, and the acute plasma proteomic response to exercise.

After one year of HIIT, participants showed highly variable fitness responses, ranging from a 28% decline to 56% increase in VO_2_ (Fig. 1A), consistent with prior reports of individual heterogeneity in trainability. To identify circulating determinants of this heterogeneity, we adopted an extreme-phenotype discovery strategy designed to maximize biological contrast rather than estimate population-level effect sizes. We selected two groups from the HIIT arm: high-responders, HR (ΔVO_2_≥10%; mean 22%; N=10 subjects) and low-responders, LR, with little or no response to training (-2%≤ΔVO_2_≤3%; N=10 subjects). Subjects from these two groups were matched on potential confounders, including sex, age, lean muscle mass, baseline peak VO_2_ (Table 1). We then profiled the circulating proteome using a DNA aptamer-based platform to quantify 1305 proteins in HR and LR subjects across 80 plasma samples (40 per group), collected at rest and after acute maximal exercise, both before training and after one year of HIIT.

**Fig. 1.**
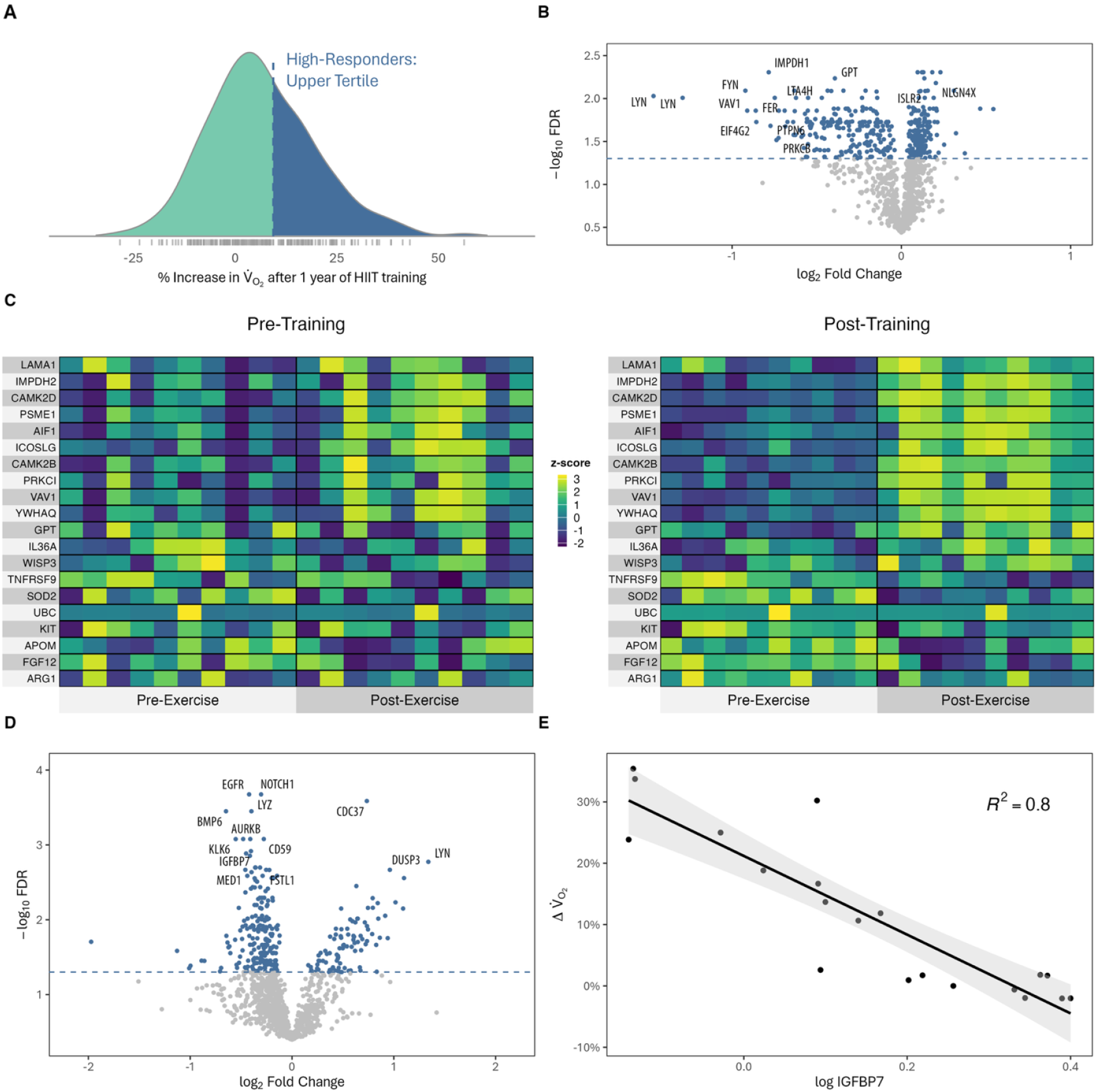
Proteomics of Trainability. (**A**) Distribution of cardiorespiratory fitness (peak VO_2_) change after one year of HIIT in 227 septuagenarian study participants. (**B**) The effect of training on the circulating proteome at rest (post-HIIT vs pre-HIIT), as measured in 10 high-responders; proteins colored in blue exhibit significant differential levels after one year of training compared to baseline (FDR < 0.05). (**C**) The effect of training on the circulating proteins before and after a bout of acute exercise in the same high-responders as in panel B. Heatmaps show the top 20 proteins whose ratio of post-to pre-exercise levels changed most significantly after training compared to baseline (FDR < 0.05). Tiles are colored by within-protein z-score scaled abundance. (**D**) Proteins differentially abundant with between high- and low-responders at rest, before training, in the discovery cohort; blue indicates significant differences (FDR<0.05). (**E**) The relationship between IGFBP7 levels measured at rest, before training, and the subsequent percent change in peak VO_2_ after one year of HIIT training, across all participants from panel D. IGFBP7 is quantified in normalized dimensionless units. HIIT indicates high-intensity interval training; FDR, false discovery rate; IGFBP7, insulin-like growth factor binding protein 7.

We first examined how acute exercise and chronic HIIT training stimulate and remodel the circulating plasma proteome, stratified by trainability. At baseline, an acute bout of maximal exercise stimulated broad proteomic shifts in both groups, with 659 proteins altered in HR individuals and 783 in LR individuals (FDR<0.05, data S1), indicating quantitatively comparable responses to physiological stress in the plasma proteome between the groups.

In contrast, chronic training produced strikingly divergent proteomic remodeling. In HR individuals, HIIT substantially altered the resting levels of 394 proteins (FDR < 0.05; Fig. 1B) and the acute exercise response of 323 proteins (post-vs. pre-exercise, FDR < 0.05; Fig. 1C). In LR individuals, however, HIIT produced no significant changes in either the resting proteome or the acute exercise response. Thus, successful physiological adaptation was mirrored by coordinated remodeling of both basal and stress-responsive proteomic states, a feature absent in low responders.

We next asked whether features of the baseline plasma proteome could predict trainability itself. At rest, prior to training, 301 proteins differed between HR and LR individuals, of which 228 predicted future fitness gains, including 95 upregulated and 133 downregulated proteins (Fig. 1D, data S1). Functional enrichment analysis revealed that the upregulated proteins in HR group are significantly enriched in pathways relevant to exercise capacity, including growth hormone synthesis, secretion and action, the VEGF signaling pathway, and cell cycle (fig. S1). No significant functional categories were found to be overrepresented in the downregulated proteins.

The circulating protein with the strongest correlation with fitness gain (R^2^=0.8, P<0.001) was insulin-like growth factor binding protein 7 (IGFBP7; Fig. 1E), a senescence-associated secretory phenotype (SASP) protein, whose baseline levels varied inversely with fitness gains. We validated this relationship using an orthogonal automated ELISA platform for IGFBP7 in 191 participants from the HIIT arm of the G100 study with available samples, confirming that lower baseline IGFBP7 levels predicted greater training-induced increases in peak VO_2_ (P=0.004, fig. S2). Training did not alter IGFBP7 levels, and the effect of training on the top 20 fitness predictors was modest, ranging from - 29% to +16%. No acute exercise–induced proteomic responses measured before training predicted future fitness gains (not shown). Together, these findings suggest that the resting plasma proteome may be primed to influence adaptive capacity, with circulating factors that could actively constrain or enhance physiological adaptation to exercise.

### IGFBP7 regulates the response to exercise training

To test whether IGFBP7 causally influences adaptive responses to exercise, we examined IGFBP7 knockout (KO) mice (*11*), beginning with baseline phenotypes that might shape training responsiveness. Compared to wild-type (WT) controls, there were no significant baseline differences in tibia length (not shown), a proxy for body size. Female KO mice exhibited modestly higher body weight (+ 9%, P=0.005), attributable to increased fat mass (+61%, P=0.002). KO mice also exhibited modestly better cardiac systolic function (Fractional Shortening [FS] 56% vs. 52%, P<0.001). In contrast, muscle mass was consistently reduced across multiple beds, including heart (14%) and gastrocnemius (15%) (Fig. 2A, fig. S3). Despite reduced muscle mass, KO mice displayed slightly better baseline exercise capacity (Fig. 2B, lower panel). The similarity in baseline fitness between KO and WT mice before training mirrored the comparable baseline fitness of HR and LR participants in the human discovery cohort.

**Figure 2.**
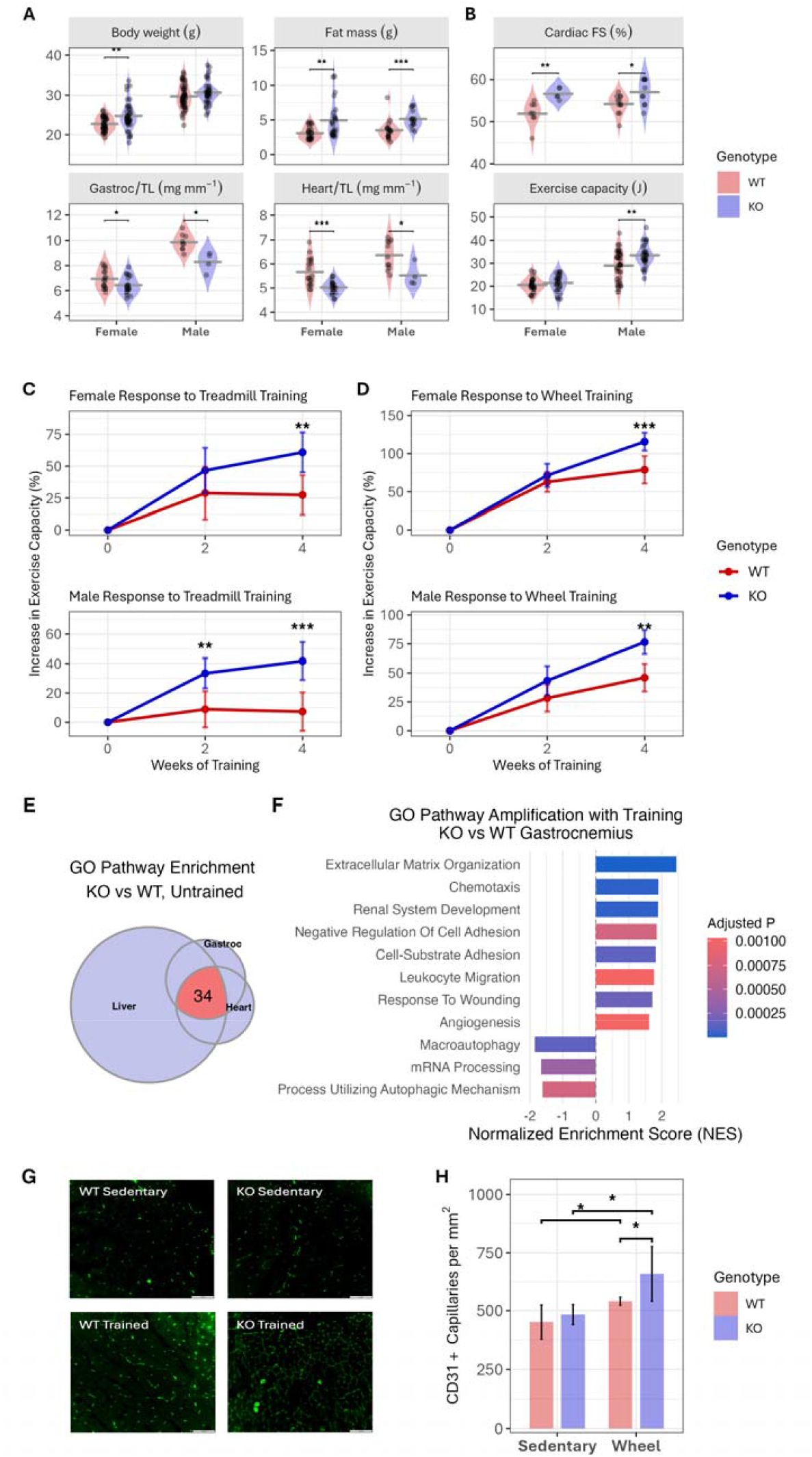
IGFBP7 deletion amplifies adaptive responses to exercise training in mice. **(A-B)** Baseline characteristics of WT and IGFBP7 KO mice before training; N=15 to 29 animals per group. **(C)** Effect of intensity-matched treadmill training on exercise capacity, stratified by sex; N=8 to 14 animals per group. **(D)** Effect of voluntary wheel training on exercise capacity, stratified by sex; N=7 to 11 animals per group. **(E)** Overlap of GO pathways positively enriched in KO vs WT at baseline; circle size is proportional to the number of enriched pathways. **(F)** GO pathways enriched in KO vs WT gastrocnemius muscle after training but not enriched at baseline. **(G)** CD31 staining of gastrocnemius sections. **(H)** Capillary density quantified as CD31+ capillaries per mm^2^ of tissue; N=4 to 5 animals per group. WT, wild-type; KO, IGFBP7 knockout; TL, tibia length; FS, fractional shortening; GO, gene ontology; ***, P<0.001; **, P<0.01; *, P<0.05.

We next tested whether loss of IGFBP7 alters the adaptive response to exercise training. Mice underwent a four-week treadmill training program (35 min sessions, 3x/week) in which the relative intensity of exercise training was comparable across animals. In particular, training intensity was set at ∼65% of each animal’s peak power output, as measured by the most recent exercise test. Peak power output was reassessed every two weeks to maintain consistent relative intensity throughout the intervention (Methods). Despite similar baseline exercise capacity, KO mice displayed markedly greater adaptation to exercise training (% increase in exercise capacity over baseline) than WT controls. After four weeks, exercise capacity increased on average 3.3-fold more in KO mice than in WT mice, with amplification observed in both sexes (2.2-fold in females and 5.7-fold in males), and the largest numeric gains seen in KO females (Fig. 2C). On an absolute scale, the change in exercise capacity and the peak exercise capacity were likewise greater in KO mice (fig. S4). Thus, IGFBP7 deletion robustly potentiated adaptation to exercise training, mirroring the inverse association between circulating IGFBP7 levels and trainability seen in humans.

We then examined the effects of IGFBP7 deletion in a distinct training model, voluntary wheel running, characterized by low intensity but high volume. Over four weeks, male and female mice ran an average of 5.4 km/day and 6.9 km/day, respectively, with no significant difference in running volume between genotypes (data not shown). This sustained running volume produced large increases in exercise capacity across sexes and genotypes, exceeding those achieved with treadmill training. Even with such a robust training effect, gains in exercise capacity remained significantly greater in IGFBP7 KO mice, surpassing those of WT controls by 54% overall (47% in females, 66% in males, Fig. 2D). This genotype-dependent difference in trainability was evident whether exercise capacity was quantified on a relative or absolute scale (fig. S4). In addition to amplifying the magnitude of adaptation, IGFBP7 deletion accelerated the kinetics of adaptation, with genotype-specific differences in training effect emerging by two weeks under both intensity- and volume-based training programs.

Finally, we tested whether increasing circulating IGFBP7 is sufficient to suppress adaptive responses to exercise training. We used an adeno-associated virus (AAV8) with a liver-specific promoter (*12*) to overexpress IGFBP7 in livers of WT and KO female mice, producing a 4-fold (P<0.001) increase in circulating IGFBP7 compared to WT levels (fig. S5A) without affecting running behavior (fig. S5B). After four weeks of voluntary wheel running, the enhanced trainability of KO mice was completely abolished by IGFBP7 overexpression (fig. S5C-D). Thus, elevating circulating IGFBP7 is sufficient to suppress exercise-induced adaptation. Notably, although IGFBP7 levels also increased in WT mice receiving the AAV-IGFBP7 vector, their training response did not decline compared with WT mice receiving the AAV-control vector, suggesting a non-linear or threshold-dependent effect of IGFBP7 on trainability.

### IGFBP7 shapes metabolic, remodeling, and angiogenic pathways

To investigate the mechanisms underlying the enhanced trainability observed with IGFBP7 deletion, we first examined training-induced changes in skeletal muscle mass. Across all muscle groups, KO mice showed an overall 9% greater increase in muscle mass with training compared to the increase in WT mice (genotype × training interaction, P<0.001), though with variation across muscle beds (fig. S6). IGFBP7 has been reported to inhibit IGF1 signaling (*13*), and to correlate inversely with circulating IGF1 levels in humans (*14*). Consistent with this relationship, circulating IGF1 levels were 40% higher in KO mice than in WT controls (P<0.01). Given the established role of IGF1 in promoting muscle growth and endurance (*15*), elevated circulating IGF1 may contribute to the amplified training-induced muscle hypertrophy observed in KO mice. Together with the observation that sedentary KO mice achieve similar or greater exercise capacity despite reduced muscle mass, these findings suggest that disproportionate muscle hypertrophy during training contributes to the enhanced adaptive response in the absence of IGFBP7.

We next asked how IGFBP7 deletion shapes molecular pathways across tissues. To isolate baseline differences independent of training, we compared gene expression profiles across genotypes in sedentary mice. Transcriptomic profiling of gastrocnemius, heart, and liver identified 11 (other than IGFBP7) differentially expressed genes (DEGs) shared across all three tissues (table S1, fig. S7A). The heart exhibited 398 differentially expressed genes, roughly four times more than gastrocnemius or liver, suggesting a greater transcriptomic impact of IGFBP7 deficiency in cardiac tissue at baseline (fig. S7A). Pathway enrichment analysis comparing untrained KO mice to WT mice revealed a striking unifying signal – all 34 GO pathways shared across tissues were positively enriched and tied to energy homeostasis and aerobic ATP generation (Fig. 2E, table S2). Of note, the oxidative phosphorylation KEGG pathway was also strongly enriched in KO mice across all tissues (fig. S7B). This is consistent with prior reports that IGFBP7 inhibits oxidative phosphorylation (OXPHOS) and mitochondrial function (*16*). These baseline differences in OXPHOS may contribute to preserved or enhanced exercise capacity in untrained KO mice despite reduced muscle mass.

Since the baseline exercise capacity of KO mice is similar to that of WT controls, the baseline transcriptomic differences may not directly account for enhanced exercise capacity. Thus, we examined transcriptional responses that were amplified by training in IGFBP7 KO compared to WT mice (Fig. 2F, fig. S8). In gastrocnemius muscle, more genes were altered by training in KO compared to WT (665 vs 403), and in the trained animals, 49 genes were differentially expressed between genotypes (fig. S8A). Notably, training induced a twofold increase in *Igf1* expression in KO mice (P < 0.001), but was unchanged in WT controls, consistent with higher plasma IGF1 levels and enhanced exercise-induced muscle growth and endurance (*15*). Further evidence of metabolic remodeling enhanced by training in the KO included positive enrichment of fatty acid metabolism and reduced mitophagy pathways (data S3). Notably, OXPHOS pathways were not further enriched with training in the KO. In contrast, training led to enrichment of OXPHOS in WT muscle, catching up with the elevated baseline levels seen in the KO animals, reinforcing that differences in OXPHOS do not account for enhanced trainability in the KO.

Pathway analysis in gastrocnemius also revealed amplification of structural remodeling programs, including pathways related to extracellular matrix (ECM) organization and angiogenesis. Multiple GO and KEGG ECM pathways were enriched in trained KO muscle compared to trained WT muscle, but not enriched at baseline (Fig. 2F, fig. S8, data S4). Of note, these included ECM remodeling that may facilitate angiogenesis, microvascular maturation, mechano-transduction, and muscle mechanics (*17-19*). Although angiogenesis-related pathways were induced by training in both genotypes, they showed significantly higher enrichment in trained KO muscle (normalized enrichment score [NES] 1.6, P = 0.001), a difference not observed at baseline (Fig. 2F). This finding is consistent with anti-angiogenic effects of IGFBP7 reported in some (*20*) but not all settings (*21*). These transcriptional findings were supported by histological analyses, which demonstrated a twofold greater training-induced increase in capillary density in KO compared with WT gastrocnemius muscle (Fig. 2F-G) but no difference at baseline. Given the central role of capillary networks in oxygen delivery and metabolic waste removal during exercise (*22*), enhanced angiogenic remodeling likely contributes to the augmented gains in exercise capacity seen with IGFBP7 deletion.

We also examined training-induced transcriptomic changes in heart and liver. In the heart, exercise training robustly upregulated pathways that promote a shift toward oxidative ATP production and substrate oxidation. In particular, both KEGG and GO pathways for tricarboxylic acid (TCA) cycle and amino acid catabolism were differentially enriched in trained KO compared to trained WT hearts, differences that were not present at baseline (fig. S9, data S4). Training in KO hearts also amplified KEGG and GO pathways of cardiac contraction and a GO pathway of sarcoplasmic reticulum calcium transport. Such effects could increase cardiac stroke volume and reserves, which could in turn enhance exercise capacity. In the liver, training-amplified responses were most prominent in pathways related to fuel oxidation pathways, including aerobic respiration, fatty acid metabolism, and the TCA cycle. These pathways were enriched by training in KO animals, but not in WT, leading to greater enrichment in trained KO livers compared to trained WT livers (fig. S10). Enhanced hepatic metabolic remodeling may support substrate availability, helping to maintain metabolic homeostasis and fuel working muscle during exercise.

Together, these data indicate that the amplified adaptive response observed in IGFBP7-deficient mice is multifactorial, impacting multiple organs essential for the mechanical and metabolic adaptation to exercise. In skeletal muscle, elevated systemic and local IGF1 signaling, enhanced metabolic remodeling, increased angiogenesis, and augmented extracellular matrix reorganization likely represent the dominant contributors to improved exercise capacity by facilitating oxygen delivery, substrate utilization, and force transmission during training. Training-induced transcriptional changes in the heart and liver may further support this phenotype by enhancing cardiac energetic capacity and substrate metabolism, respectively. Prior work showing that isolated improvements in cardiac contractile performance may not increase exercise capacity (*23*) suggests these cardiac adaptations may not be the primary drivers of an enhanced training response in mice. However, they could contribute and be important clinically if parallel changes occur in humans. The coordinated remodeling of skeletal muscle metabolic, vascular, and structural programs appears to be the principal mechanism through which IGFBP7 deletion amplifies exercise-induced gains in exercise capacity. Collectively, these convergent adaptations provide a mechanistic framework for the amplified gains in exercise capacity observed with IGFBP7 deletion.

### IGFBP7 levels correlate with multiple human health outcomes

Given the close relationship between cardiorespiratory fitness and health, we asked whether IGFBP7 levels are associated with incident disease across organ systems in humans. We analyzed 52,033 UK Biobank participants with baseline proteomic measurements (Olink; collected 2006–2010) and a median follow-up of 16.3 years, fitting age- and sex-adjusted proportional hazards models for seven incident outcomes: mortality, heart failure, chronic obstructive pulmonary disease, cirrhosis, type 2 diabetes, chronic kidney disease, and cancer. Circulating IGFPB7 levels increased with age in both men and women (fig. S11A). Nevertheless, even after adjustment for age and sex, IGFBP7 showed strikingly similar dose–response associations across six of the seven outcomes (all P < 10^-16^), with lower levels consistently associated with reduced risk (Fig. 3A). Cancer risk differed from this pattern, with no excess risk at low to intermediate IGFBP7 levels and a modest increase in hazard observed only at higher levels (fig. S11B). Hazard curves for these six outcomes crossed 1.0 within a narrow IGFBP7 range (0.5–1 SD units), allowing stratification into risk groups. Event rates were consistently higher in the upper quartile, with 10-year cumulative risk that was 2.9–11 times greater than in the lowest quartile (Fig. 3B). Together, these findings indicate that circulating IGFBP7 levels are associated with long-term risk across multiple age-related diseases, paralleling the breadth of health outcomes linked to cardiorespiratory fitness.

**Figure 3.**
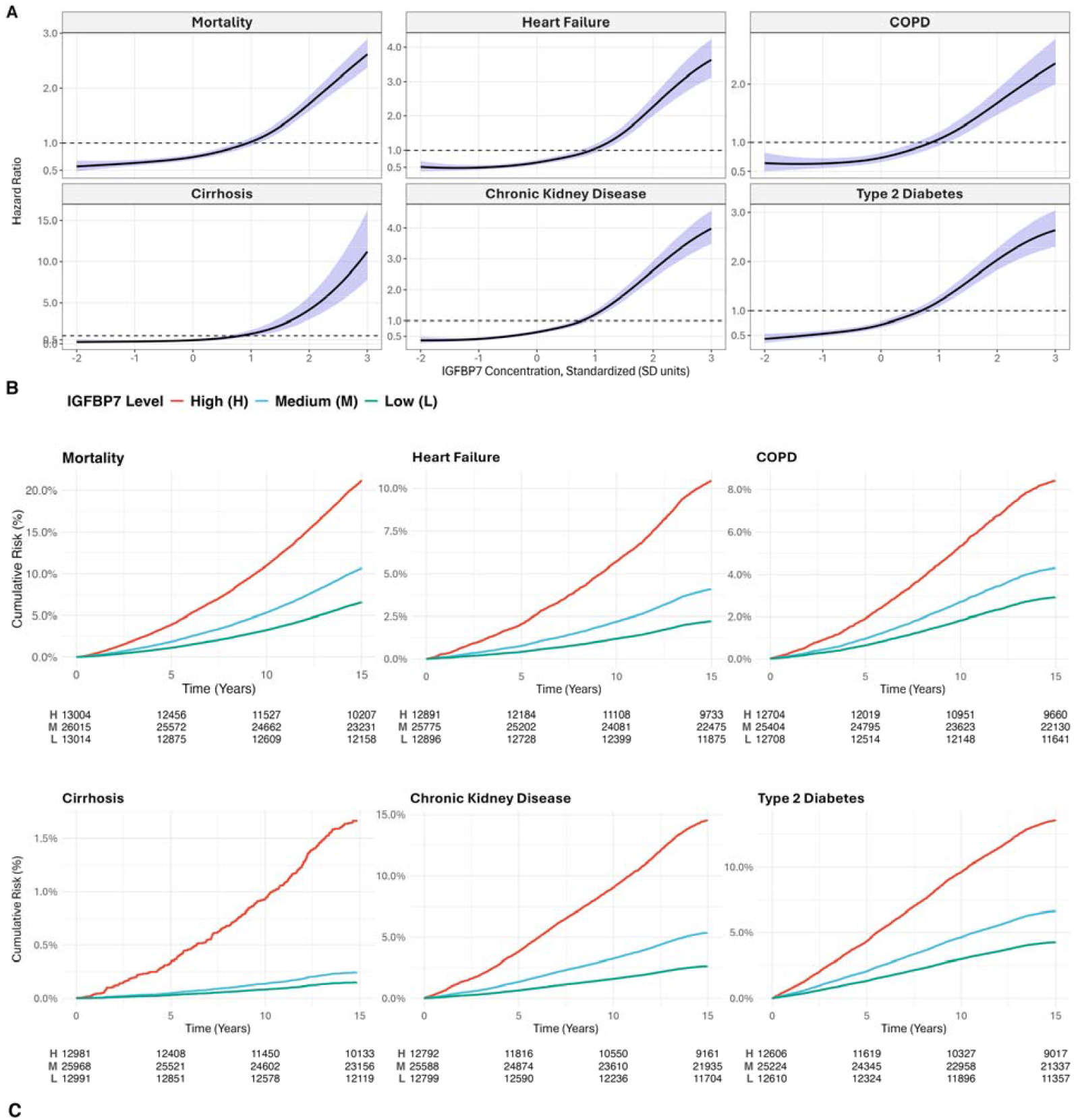
Circulating IGFBP7 levels are associated with age-related health outcomes across multiple organ systems. **(A)** Relationship between circulating IGFBP7 levels and mortality or incident disease in age- and sex-adjusted proportional hazards model fitted to participant data from the UK Biobank. IGFBP7 was measured by the Olink platform. **(B)** Cumulative incidence curves for patient outcomes modeled in panel A, with IGFBP7 categorized as Low (lowest quartile), Medium (interquartile range), or High (highest quartile). At-risk tables reflect raw counts. **(C)** Conceptual model summarizing proposed relationships among IGFBP7, exercise training, adaptive capacity, and health. Elevated IGFBP7 constrains physiological adaptation to exercise, limiting gains in fitness, which may adversely influence health outcomes. IGFBP7 may also affect health through exercise-independent pathways. COPD indicates chronic obstructive pulmonary disease.

## Discussion

By integrating human proteomics, genetic manipulation in mice, and large-scale epidemiologic data, we identify IGFBP7 as a circulating regulator of adaptation to exercise. In older adults undergoing high-intensity interval training, lower baseline IGFBP7 levels predicted greater improvements in cardiorespiratory fitness and were associated with reduced age- and sex-adjusted mortality and incident disease across multiple organ systems in the UK Biobank, a large prospective cohort. In mice, genetic deletion of IGFBP7 amplified exercise-induced gains in exercise capacity, whereas somatic overexpression abolished this advantage, demonstrating that circulating IGFBP7 levels can constrain physiological adaptation in adulthood. The increase in IGFBP7 levels with age in humans (fig. S11A), could contribute to age-related declines in adaptation. Given IGFBP7’s known role as a senescence factor (*24*), these findings implicate IGFBP7 as a molecular brake on trainability, linking aging biology to interindividual differences in exercise responsiveness and, potentially, to broader health outcomes.

Our results also illuminate the biology of trainability, a clinically important form of physiological adaptation. Acute exercise elicited similar widespread changes in the circulating proteome in both high- and low-responders, none of which predicted future adaptation. Instead, baseline features of the resting proteome strongly predicted training responsiveness, supporting the concept that adaptive capacity is preconfigured before training begins. Among more than two hundred baseline predictors, IGFBP7 emerged as the dominant predictor of trainability in our older human cohort, consistent with its established role as a senescence-associated secretory phenotype factor whose circulating levels increase with age and disease (*25, 26*).

IGFBP7 is a secreted glycoprotein that circulates systemically and localizes to the extracellular matrix (ECM), positioning it to influence multiple processes relevant to aging and adaptation (*21*). Prior work implicates IGFBP7 in cellular senescence, satellite cell proliferation, ECM remodeling, angiogenesis, mitochondrial metabolism, and insulin/IGF-1 signaling (*13, 21, 27-29*). Our transcriptional profiling and structural analyses support a role for many of these pathways in shaping the enhanced adaptive response observed with IGFBP7 deletion. At baseline, mitochondrial oxidative pathways were enriched in IGFBP7-deficient muscle, likely contributing to preserved or modestly enhanced exercise capacity despite reduced muscle mass. With training, IGFBP7-deficient mice exhibited exaggerated hypertrophy, enhanced induction of *Igf1*, amplification of lipid metabolic programs, and a marked increase in capillary density, accompanied by upregulation of angiogenic and extracellular matrix remodeling pathways. Notably, an enhanced microvascular response has also been reported to distinguish animals genetically selected for high versus low training responsiveness, underscoring the relevance of vascular remodeling to adaptive capacity (*30*). Together, these findings support a model in which IGFBP7 restrains both metabolic and structural axes of adaptation, limiting the extent to which tissues remodel in response to repeated physiological stress.

In addition to fitness, IGFBP7 levels were associated with a broad spectrum of age-related health outcomes in the UK biobank, including heart failure, chronic kidney disease, type 2 diabetes, and all-cause mortality. Two broad explanations seem plausible (Fig. 3C). First, the health benefits of enhanced trainability could be mediated through improved fitness. Amplification of training-induced fitness would be expected to confer well-established benefits for cardiovascular and systemic health. However, enhanced trainability may also induce adaptations that do not directly impact fitness but increase resilience to disease. Consistent with this possibility, both the resting circulating proteome and the acute proteomic response to exercise exhibited numerous changes in high responders, whereas no such changes were observed in low responders (Fig. 1B,C). These proteomic adaptations may mark or mediate broader physiological resilience relevant to disease. A second possibility is that enhanced trainability itself, independent of exercise, confers resistance to disease. Trainability is an archetypal form of physiological plasticity, and a system primed to respond robustly to stress may be better able to withstand pathological insults. Notably, some of the physiologic stresses imposed by exercise, such as thermal, ionic, metabolic, or hemodynamic disturbances, also arise in disease. Such a framework suggests that modulating IGFBP7 could enhance organismal resilience even in sedentary individuals, a concept supported by prior preclinical studies (*31*). These mechanisms are not mutually exclusive and together suggest that constrained physiological plasticity may link aging biology to both diminished exercise responsiveness and increased disease risk.

Several limitations merit consideration. Although we identified 228 proteins at baseline that predicted fitness responses, we only validated the functional role of the strongest predictor, IGFBP7. Given that our targeted proteomics platform captured only a subset of the circulating proteome, other unmeasured proteins likely also contribute to exercise adaptation as either constraints or enablers. Investigating these candidates to more fully map the regulatory landscape of exercise adaptation will be important in future studies. We did not directly test whether pharmacological inhibition of IGFBP7 in adults enhances trainability, although the effects of somatic overexpression suggest adults retain plasticity in the training response. The value of IGFBP7 inhibition for health outcomes or to improve athletic performance remains to be determined. Safety is a concern; IGFBP7 knockout mice develop liver tumors (*11*), raising the possibility that this constraint on adaptation may also serve to limit neoplastic transformation. Reassuringly, we observed no cancer signal with low IGFBP7 in the UK Biobank cohort. Notably, prior reports of tumorigenesis were in sedentary, caged animals, and exercise is reported to reduce cancer risk (*32, 33*), so the cumulative effects may differ in active individuals. Nevertheless, both the benefits and risks associated with IGFBP7 inhibition would need to be rigorously evaluated before considering therapeutic interventions.

In summary, our work identifies IGFBP7 as a genuine “trainability gene”: its loss does not substantively alter baseline fitness, but it primes the organism for superior adaptation to training. In older adults, where declining adaptability contributes to loss of function and increased disease risk, understanding and potentially modulating this pathway may offer new opportunities to enhance the benefits of physical activity and impact health outcomes. Future studies will be required to determine whether this biology can be safely and effectively harnessed in humans.

## Data Availability

All data, code, and materials used in the analysis will be made available to interested researchers upon request and completion of the standard institutional materials transfer agreements (MTAs) as well as payment of applicable shipping or transfer costs. RNA sequencing data will be deposited in a public database, along with relevant metadata once manuscript is accepted for publication.

## Acknowledgments

This research has been conducted using the UK Biobank Resource under Application Number 197947. We thank the NeXt Move Core Facility, Norwegian University of Science and Technology (NTNU), and the Clinical Research Facility at St. Olav Hospital for excellent assistance during the testing periods in the Gen100 study. AI-assisted technologies were used to check grammar and editing and to generate the images in Figure 3C. The study was approved by the Regional Committee for Medical Research Ethics (REC Number 2016/1874).

## Funding

The funding organizations are listed below; they had no role in the design and conduct of the study; in the collection, analysis, and interpretation of the data; or in the preparation, review, or approval of the manuscript.

National Institutes of Health grant R35 HL155318 (AR)

National Institutes of Health grant R01HL173129-01 (NH)

National Institutes of Health grants R01HL171201 and R01HL169272 (HL)

National Institutes of Health grant R03 HL177119 (JRh)

American Heart Association grants 23MERIT1038415 and SFRN (URLs: https://doi.org/10.58275/ AHA.24SFRNPCN1284382.pc.gr.194135, https://doi.org/10.58275/ AHA.24SFRNCCN1276092. pc.gr.194131, and https://doi.org/10.58275/AHA.25ISFRNCP1501445.pc.gr.231914 (AR)

American Heart Association grant 24SCEFIA1253853 (HL)

Challenger Foundation Career Development Award (QZ)

Research Council of Norway (UW, DS)

K.G. Jebsen Foundation for Medical Research, Norway (UW, DSt)

Norwegian University of Science and Technology (NTNU) (UW, DSt)

Central Norway Regional Health Authority, St Olav’s Hospital, Norway (UW, DSt)

National Association for Public Health, Norway (UW, DSt)

## Author contributions

Conceptualization: AR, NH, JR

Data Curation: NH, BL, QZ, CW

Formal Analysis: NH, BL, QZ, CW, JG

Methodology: NH, QZ, HL

Investigation: NH, QZ, YC, MQ, AK, SM, VC, GG, SW, DS, JRh, JSG,

Funding acquisition: AR, UW, DSt, HL

Project administration: AR, NH Resources: UW, DSt, HL

Supervision: AR

Visualization: NH, BL, CW, AR

Writing – original draft: NH, AR

Writing – review & editing: NH, AR, HL, BL, ABL, JR, UW, DS

## Supplementary Materials

Materials and Methods

Tables S1 to S2

Figs. S1 to S11

Data S1 to S4

