## Supplementary Methods, Tables, and Figures for "Circulating Senescence Protein Links Exercise Adaptation to Health Outcomes"

**The PDF file includes:**

Materials and Methods

Figs. S1 to S11

Tables S1 to S2

**Other Supplementary Materials for this manuscript include the following:**

Data S1 to S4

### Materials and Methods

**Human Studies**

### Generation 100 Study Participants

The Generation 100 (G100) study design and primary outcomes have been previously reported *(9)*. The present analysis focused on participants assigned to the high-intensity interval training (HIIT) arm. For plasma proteomic analyses, we selected a discovery cohort of 20 individuals representing distinct extremes of trainability. Ten high-responders (HR) were defined by a robust improvement in cardiorespiratory fitness after one year of HIIT (relative increase in peak VO_2_ >10%). Ten low-responders (LR) were defined by minimal fitness change (–2% to +3%); individuals with marked fitness declines were excluded to minimize confounding by occult illness or major comorbidity. Groups were matched for baseline peak VO_2_ (absolute and peak predicted), lean muscle mass, age, height, sex (all male), and training adherence as documented in training questionairres. Peak predicted VO_2_ was adjusted for age, sex, and height. To validate associations between IGFBP7 and trainability, we analyzed all participants from the HIIT arm with available baseline plasma samples who completed both baseline and one-year post-training peak VO_2_ testing.

### Proteomic profiling and analysis

### Venous blood samples were collected immediately before and after treadmill VO_2_ testing in EDTA tubes, processed and stored as previously reported (9). Relative plasma protein abundance was quantified using the SomaScan aptamer-based platform (SomaLogic), measuring 1305 proteins in raw fluorescence units (RFU). RFUs were normalized using the median of on-plate calibration samples for each analyte, and analyses were restricted to proteins with a coefficient of variation <15% across plates. Normalized RFUs were log-transformed for statistical analyses. Effect sizes were estimated as differences in log-transformed abundance and reported as log₂ fold changes.

### *Study design*

The study used a repeated-measures exercise design incorporating three factors: 1). Acute exercise status: pre- vs post-exercise within a visit; 2) Training status: pre-training (baseline) vs post-training; 3) Trainability group: HR vs LR, defined by one-year change in peak VO_2_. Training-induced change in cardiorespiratory fitness was summarized as the percent change in peak oxygen uptake (VO₂). This phenotype was used as a continuous measure of trainability in regression analyses.

### *Statistical analysis*

### Proteomic analyses were conducted at the protein level. Primary comparisons included: 1) Acute exercise effects at baseline; 2) Training-induced modifications of the acute exercise response (change in the post/pre response after training; “ratio-of-ratios”); 3) Training effects on the resting proteome (post- vs pre-training at rest); 4) Baseline differences between HR and LR at rest; 5) Associations between baseline protein levels and training-induced VO₂ change. Within-participant contrasts were assessed using paired t-tests on log-transformed RFUs, stratified by trainability group when appropriate. Baseline HR–LR comparisons used two-sample t-tests. Associations between baseline protein abundance and fitness gain were assessed using linear regression models. Multiple testing was controlled using the qvalue procedure (false discovery rate 0.05). All analyses were performed in R.

**IGFBP7 ELISA measurements and analysis**

Plasma IGFBP7 concentrations were quantified using the Ella automated immunoassay platform (ProteinSimple) with the Simple Plex Human IGFBP-rp1/IGFBP-7 Cartridge (SPCKB-PS-003329, 72 samples per plate). Frozen plasma samples were thawed on ice and then diluted 1:100 in the manufacturer’s sample diluent before loading onto the cartridge. Assays were performed according to the manufacturer’s instructions, and analyte concentrations were calculated from manufacturer supplied standard curves specific to each lot of cartridges. Batch effects across assay runs were corrected using an empirical Bayes linear model (limma R package) with sex as a covariate; outliers with extreme standardized IGFBP7 values (< −4 or > 5 SD) were excluded. The training effect, defined as the percent increase in peak VO₂, was modeled as a linear regression on age, sex, baseline VO₂ and baseline IGFBP7. All analyses were conducted in R.

### UK Biobank Analysis

Baseline plasma IGFBP7 levels were measured as part of the UK Biobank Pharma Proteomics Project (UKB-PPP) using the Olink Explore 3072 proximity extension assay (PEA), providing normalized protein expression (NPX) values, as described <https://biobank.ndph.ox.ac.uk/showcase/refer.cgi?id=4654>). Health outcomes were derived from linked hospital, primary care, cancer registry, and death records. Participants were followed from baseline until incident outcome or administrative censoring (August 1, 2025). We screened 27 incident health outcomes for an association with baseline IGFBP7 levels and selected seven outcomes for their health impact and organ diversity, including death, heart failure, chronic obstructive pulmonary disease, chronic kidney disease, liver cirrhosis, type 2 diabetes, and all-cause cancer. Time-to-event (measured in days) associations were evaluated using Cox proportional hazards models adjusted for age and sex. IGFBP7 was standardized and modeled using penalized splines for continuous analyses. For categorical analyses, IGFBP7 was grouped into low (≤25^th^ percentile), medium (25^th^-75^th^ percentile), and high (≥75^th^ percentile) categories. Adjusted Cox curves and adjusted cumulative incidence estimates were generated using standard methods. P-values for the associations between IGFBP7 and outcomes were assessed with Bonferroni-correction to account for the initial screen of 27 outcomes.

**Animal Studies**

Wild-type C57BL/6J (000664) mice were purchased from Jackson Laboratory. IGFBP7 knockout mice on a C57BL/6J background were provided by Dr. Devanand Sarkar. All animal experiments were performed with mice aged 3-6 months unless otherwise stated. Animals were anesthetized with ketamine and xylazine prior to sacrifice and tissue collection. All animal studies were approved by the Institutional Animal Care and Use Committees at Massachusetts General Hospital and the University of Michigan.

### IGFBP7 Overexpression

AAV8-mediated IGFBP7 overexpression was achieved via tail-vein injection of AAV8-TBG-IGFBP7 (2 × 10¹¹ TU per mouse) in adult female mice one week before voluntary wheel training. After four weeks of training, hepatic IGFBP7 expression was quantified by RT-qPCR and circulating IGFBP7 by ELISA (Abcam 245712).

### IGFBP7 and 18S qPCR Primers:

| Name | Forward | Reverse |
| --- | --- | --- |
| mIGFBP7 | CTGGTGCCAAGGTGTTCTTGA | CTCCAGAGTGATCCCTTTTTACC |
| m/r-RPS18 | CATGCAGAACCCACGACAGTA | CCTCACGCAGCTTGTTGTCTA |

### Mouse Echocardiography and Body Composition

Transthoracic echocardiography was performed on unanesthetized mice using an E90 system (GE Healthcare) equipped with an L8-18i-D (16 mHz) transducer. Fractional shortening as well as LV cavity and wall thickness dimensions were made from M-mode parasternal short-axis images at the mid-papillary level. Body composition was evaluated in unanesthetized animals by quantitative nuclear magnetic resonance spectroscopy (EchoMRI analyzer; Echo Medical Systems, Houston, TX), yielding estimates of lean and fat body mass.

### Exercise Testing and Training

***Exercise Testing***

Exercise testing was assessed using a 6-lane motorized treadmill (Columbus Instruments). Following acclimatization, mice underwent a continuous power ramp protocol to exhaustion. Exercise capacity was quantified as total mechanical work performed.

All acclimatization, testing, and treadmill training were performed with the treadmill tilted at a fixed angle of 20 degrees. The angle was chosen to enable an animal to achieve higher exercise intensity (power output) at lower treadmill velocities. Exercise testing was performed during morning hours (8am-12pm) in a temperature controlled room at 70F.

Mice were acclimatized to the treadmill over 3 days. On day 1 they ran for 5 min at a velocity of 5m/min and on day 2 for 5 min at 10m/min. On day 3, mice were acclimated to faster speeds by running them for 10 min, starting at 10m/min and increasing by 2m/min^2^ until a speed of 30m/min was reached, or exhaustion was reached, whichever occurred first. Acclimatization was performed with up to 6 mice (1 per lane) simultaneously.

Exercise testing was performed on individual mice. The protocol consisted of two phases, a warmup and a run to exhaustion, in which mice ran at a progressively increasing exercise intensity. Intensity was defined as the power output (milliwatts, mW) required for the mouse to keep pace with the treadmill. The 5-minute warmup began at an intensity of 5mW and ended at 10mW, increasing continuously at a rate of 1mW/min. The run to exhaustion began at an intensity of 10mW and increased continuously (3mW/min), similar to the power ramps used in cycle ergometry to measure human exercise capacity. The treadmill velocity during both the warmup and the run to exhaustion was tailored to elicit the desired power ramp from each animal. The velocity needed to elicit a specific power output was calculated for a given mouse by solving the following equation: P = v*m*g*sin(20 degrees)/60 (P = power in mW, v=treadmill velocity in m/min, m = mouse mass in grams, g = acceleration due to gravity = 9.8 m/s^2^). A power ramp of 3mW/min corresponded approximately to an acceleration of 2m/min^2^ and a duration on the ramp of 10-25 min depending on the animal’s fitness. Each mouse was motivated to run by tapping its tail or blowing compressed air whenever it descended more than half way down the length of the treadmill towards the electrical stimulus pad. Exhaustion was defined as occurring when the mouse could not keep pace with the treadmill for more than 3 seconds without descending all the way to the stimulus pad, and moreover failing this task 3 times in a row. Exhaustion was corroborated by point of care lactate testing (Nova Biomedical Lactate Plus) from a nick of the tail tip within 10s of exhaustion; animals routinely achieved peak lactate values of 10-15mM (resting lactate of 1-2mM).

Exercise capacity was quantified as the total amount of mechanical work completed by the mouse by the time exhaustion was reached: W = d*m*g*sin(20 degrees)/1000 (W = work in Joules, d = total distance run in meters, m = mouse mass in grams, g = acceleration due to gravity = 9.8m/s^2^).

***Exercise Training: Treadmill***

Before treadmill training, baseline exercise capacity was measured in every animal. Animals were subsequently trained three times a week (Mon, Wed, Fri) for 4 weeks. Each exercise session consisted of a 5 min warmup followed by a 30 min run at constant relative intensity, as measured by power output. Warmup started at a treadmill velocity of 5m/min (20 degree angle, as with testing) and over 5 minutes it increased to the target velocity for the exercise session. The 30 min training run was performed at a *matched* relative intensity across all animals. Each animal trained at a target treadmill velocity that elicited ~65% of the peak power it achieved on the baseline exercise test. This target intensity fell below the mouse’s critical power, enabling every mouse to complete the 30-minute running session without failing early due to exhaustion. Moreover, using relative intensity as the target enabled all animals to fully complete the same training dose despite a range of baseline fitness and body weights. Completing all 12 sessions of the training protocol was a prerequisite to be included in the analysis.

To improve the throughput of training, animals were trained in groups of up to 3 animals that were well-matched. In particular, the (single) treadmill velocity for the group session had to elicit a power output that fell in a range of 60-70% of each group member’s peak power output. Tail tapping and puffs of compressed air were used to motivate animals to run, just as with exercise testing as described above.

To compensate for fitness adaptation and thereby maintain a similar running intensity throughout training, exercise capacity was remeasured in all animals after two weeks of training (on Saturday and Sunday). Treadmill velocities for each animal’s exercise sessions were then recalculated to achieve the target intensity (65% of peak power) for the final two weeks of training. Following 4 weeks of training each animal underwent a 3^rd^ exercise test to measure exercise capacity.

Trainability was quantified both as a relative fitness change, percent increase in exercise capacity over baseline, as well as an absolute fitness change, the difference between post- and pre-training exercise capacity.

***Exercise Training: Wheels***

Before wheel training baseline exercise capacity was measured in every animal. Each animal was then individually housed in a cage containing a Starr Life running wheel (4.5 inch diameter); wheel revolutions per 15 seconds were monitored electronically for the duration of the 4-week training period. Exercise capacity was measured after 2 weeks of running and after completion of the full training period. Trainability was quantified as above.

### Capillary Density Measurements

Frozen gastrocnemius muscle samples were embedded in OCT and cryosectioned at 7 μm thickness using a cryostat. Sections were air-dried, fixed in 4% paraformaldehyde for 10 min, permeabilized with 0.2% Triton X-100, and blocked with 5% BSA. Endothelial cells were detected by immunofluorescence using an anti-CD31 primary antibody (Abcam, ab222783, diluted 1:150) incubated overnight at 4°C, followed by a fluorophore-conjugated secondary antibody (Fisher Scientific, A21206, diluted 1:200) for 1 h. Images were acquired using Leica Thunder Imager under identical exposure settings, and CD31-positive vessel density per mm^2^ quantified using ImageJ with consistent thresholding parameters.

### Tissue Transcriptomics

***Voluntary wheel running and tissue collection***.

Individually housed female mice were subjected to voluntary wheel running for four consecutive weeks using running wheels (STARR Life Sciences). After completion of the running protocol, mice were rested for 2 days, and heart, gastrocnemius, and liver tissues were harvested, snap-frozen in liquid nitrogen, and stored at −80 °C until processing.

***RNA extraction and RNA-seq.***

Total RNA was isolated from heart, gastrocnemius, and liver tissues using the RNeasy Plus Universal Mini Kit (QIAGEN, Cat. No. 73404) according to the manufacturer’s instructions. mRNA-seq libraries were prepared by Novogene using poly(A) enrichment, followed by paired-end sequencing (2 × 150 bp) on an Illumina NovaSeq X Plus platform (Illumina, USA).

***RNA-seq bioinformatic analysis.***

Raw RNA-seq reads (FASTQ) were filtered by Novogene to remove adapter-contaminated, poly-N–containing, and low-quality reads. Clean paired-end reads were aligned to the reference genome using HISAT2 v2.0.5 (with gene model annotation), and gene-level raw counts were generated with featureCounts v1.5.0-p3. Raw gene‑level counts were QC’d, and analyzed separately for gastrocnemius, heart, and liver with sample metadata (genotype, exercise, cohort/location); batch effects were assessed by principal components analysis and gastrocnemius counts were adjusted for location where needed using Combat-Ref. Differential gene expression was fit with DESeq2 negative‑binomial GLMs, and results were augmented with limma‑voom moderated t‑statistics for gene ranking. Pathway enrichment used clusterProfiler GSEA (GO and KEGG pathways; Benjamini-Hochberg FDR < 0.05) after Ensembl‑to‑Entrez mapping. We defined four criteria by which to infer pathway amplification in trained KO mice. These amplification types included: (1) pathways enriched by training in KO but not WT, (2) pathways enriched in trained KO vs trained WT and concordantly enriched by training in KO (training‑responsive), (3) pathways enriched in trained KO vs trained WT but not enriched at baseline (sedentary KO vs WT), and (4) interaction‑based enrichment by fitting a genotype×exercise model, ranking genes by the moderated t‑statistic for the interaction term (genotype‑specific training effect), and performing GSEA on this ranked list.

### Muscle mass analysis

Training-induced hypertrophy was assessed using linear mixed-effects models using the lme4 package in R with genotype, training status, and sex as fixed effects and tissue as a random effect. Log-transformed tissue weights (normalized to tibia length) were modeled as a function of genotype, training status (sedentary vs. wheel), and sex, including a genotype-by-training interaction term. To quantify the global differential impact of genotype on training-induced hypertrophy, we exponentiated the genotype-by-training interaction coefficient and reported it as the percent change in the training effect in knockout compared to wild-type mice.

**Statistical Analysis**

For analyses of mouse phenotypes, continuous measures were summarized as means and 95% confidence intervals where indicated. Comparisons between genotypes or training conditions were evaluated primarily with two‑tailed, unpaired Student’s t‑tests. For change‑from‑baseline measures where distributional assumptions were uncertain, nonparametric Wilcoxon rank‑sum tests were applied. Statistical significance was assessed at a two‑sided α = 0.05.

### Supplementary Tables

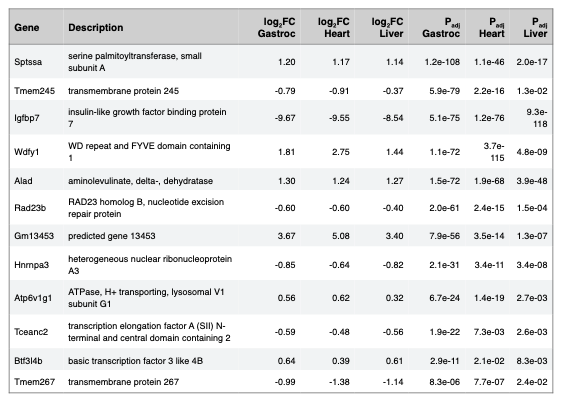

**Table S1.** **Differentially expressed genes shared across untrained mouse tissues**. Up and downregulated genes in KO vs WT, shared across gastrocnemius, heart, and liver (FDR < 0.05). Genes with |log_2_FC| > 0.5 in at least one tissue are displayed. FC indicates fold-change; P_adj_, adjusted p-value; Gastroc, gastrocnemius.

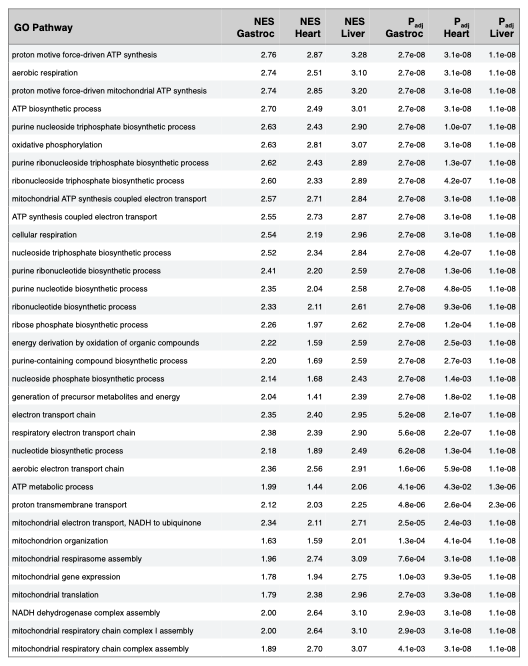

**Table S2. Differentially enriched pathways shared across untrained mouse tissues.** Positively enriched GO biological process (BP) pathways in KO vs WT at baseline, shared across gastrocnemius, heart, and liver (P_adj_ < 0.05); no negatively enriched pathways were shared. GO indicates Gene Ontology; NES, normalized enrichment score; P_adj_, adjusted p-value; Gastroc, gastrocnemius.

**Table S2.** **Differentially enriched pathways shared across untrained mouse tissues**. Positively enriched GO biological process (BP) pathways in KO vs WT at baseline, shared across gastrocnemius, heart, and liver (FDR < 0.05); no negatively enriched pathways were shared. GO indicates Gene Ontology; NES, normalized enrichment score; Padj, adjusted p-value; Gastroc, gastrocnemius.

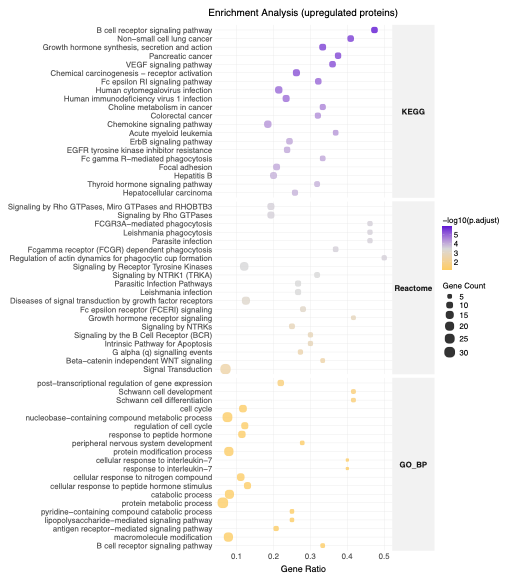

### Supplementary Figures

**Figure S1. Positively enriched pathways in High Responders at baseline.** Enrichment analysis performed from differential protein abundance in High- vs Low- Responders before training. Top 20 significantly biological terms from each category are shown (FDR < 0.1).

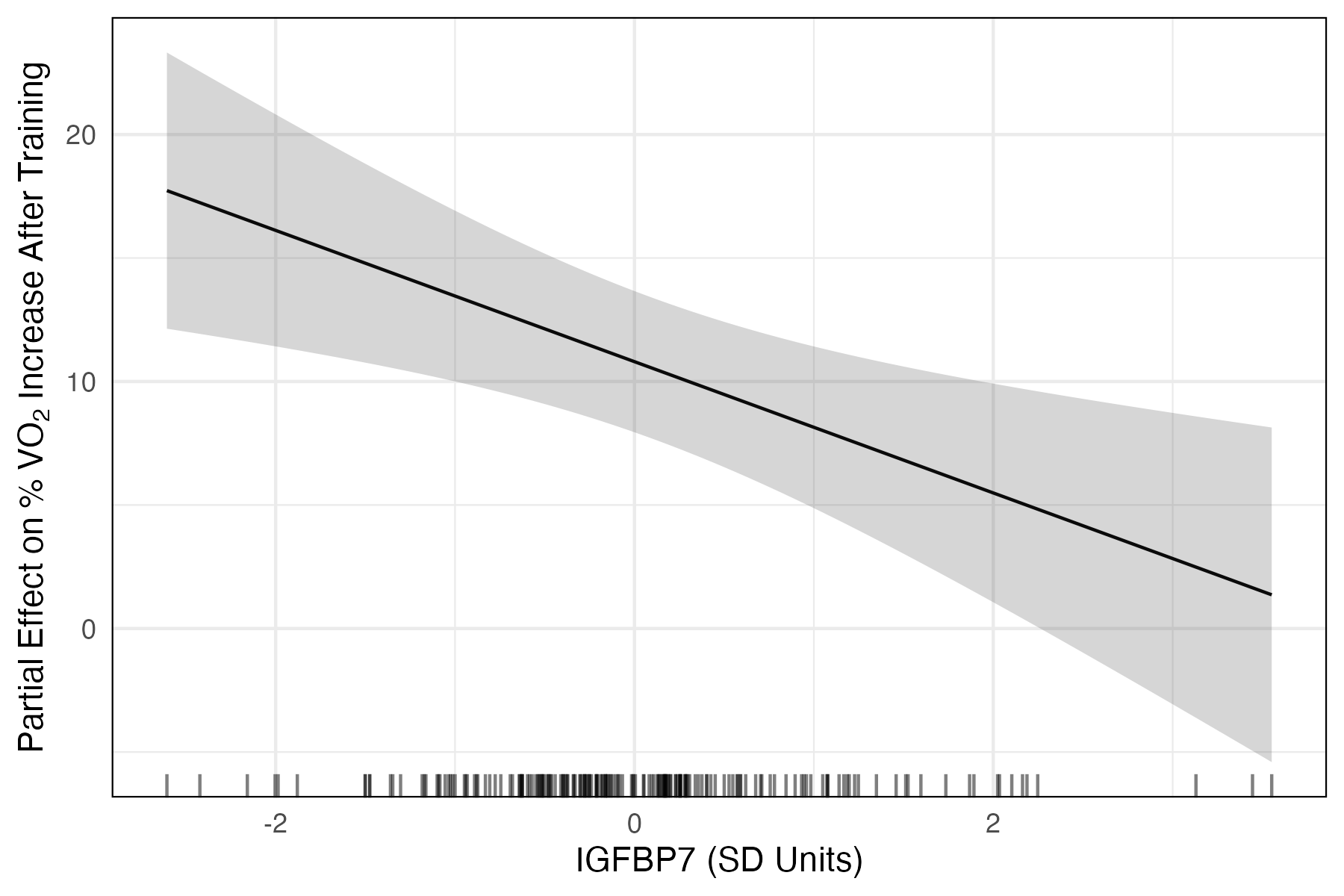

**Figure S2. Baseline IGFBP7 as a predictor of fitness gain.** Fitness gain was quantified as the % change in peak VO_2_ from baseline to 1 year of exercise training. Fitness gain was regressed on age, sex, baseline VO_2_, and IGFBP7 levels as measured by ELISA, in N=191 participants from the HIIT arm of the G100 study. Only exercise tests with peak RER ≥ 1.05 were included to increase the likelihood that peak VO_2_ reflected maximal effort. The graph depicts the relationship between baseline IGFBP7 levels prior to training with % fitness improvement after one year of training; results shown for males, with age and baseline VO_2_ held at mean values. Error bands reflect 95% confidence intervals. P-value for the IGFBP7 beta coefficient was 0.004. VO_2_ indicates oxygen consumption in L/min; HIIT, high-intensity interval training; G100, generation 100; RER, respiratory exchange ratio.

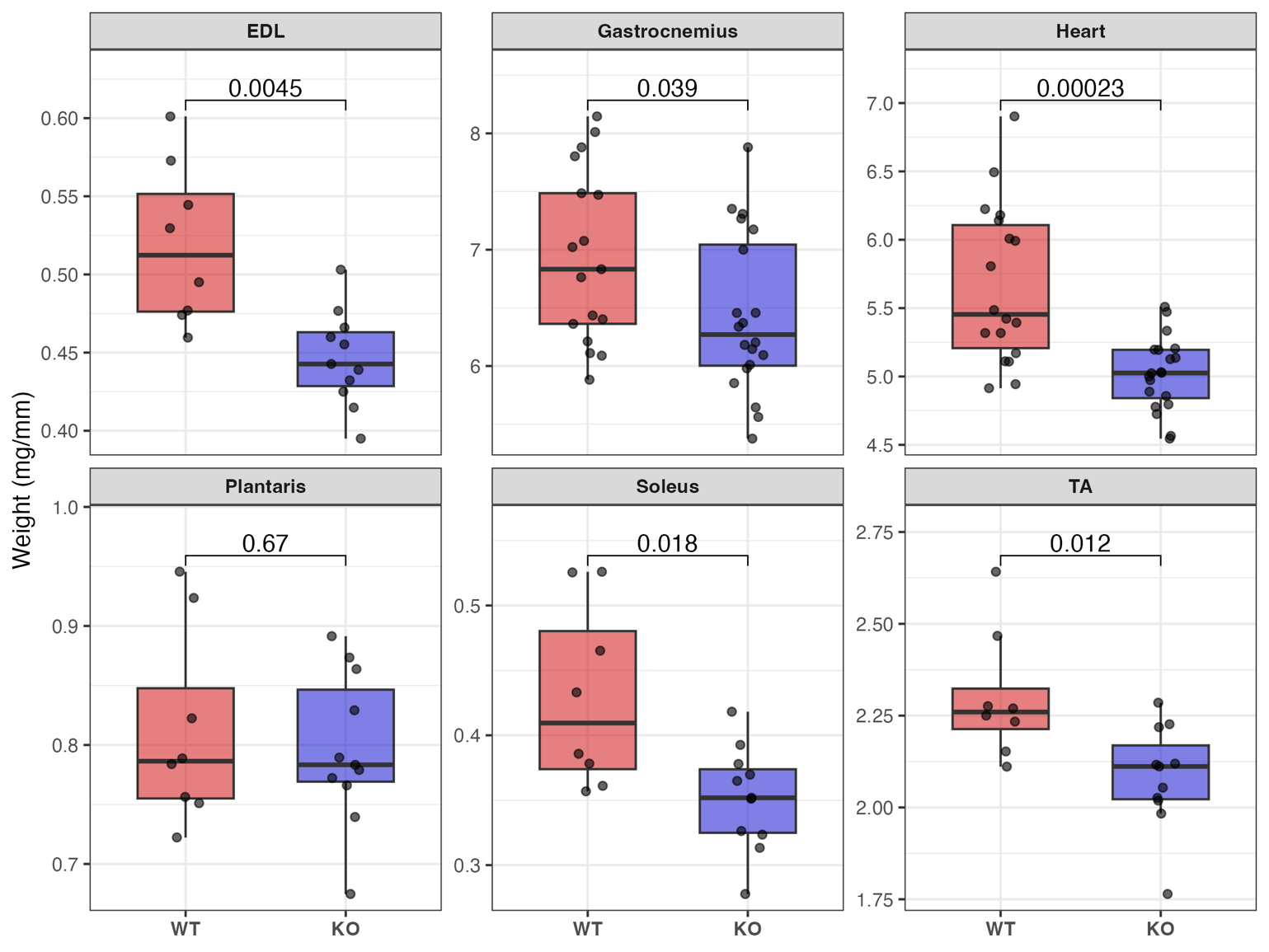

**Figure S3. Female muscle weights at baseline.** Comparison of muscle weights between IGFBP7 knockout (KO, blue boxes) and wildtype (WT, red boxes) female mice that were untrained. Muscle weight was normalized to tibia length. P values for the differences in muscle weights are annotated on the cross bars. Error bars indicate 95% confidence intervals. EDL indicates extensor digitorum longus; TA, tibialis anterior; WT, wild-type; KO, IGFBP7 knockout.

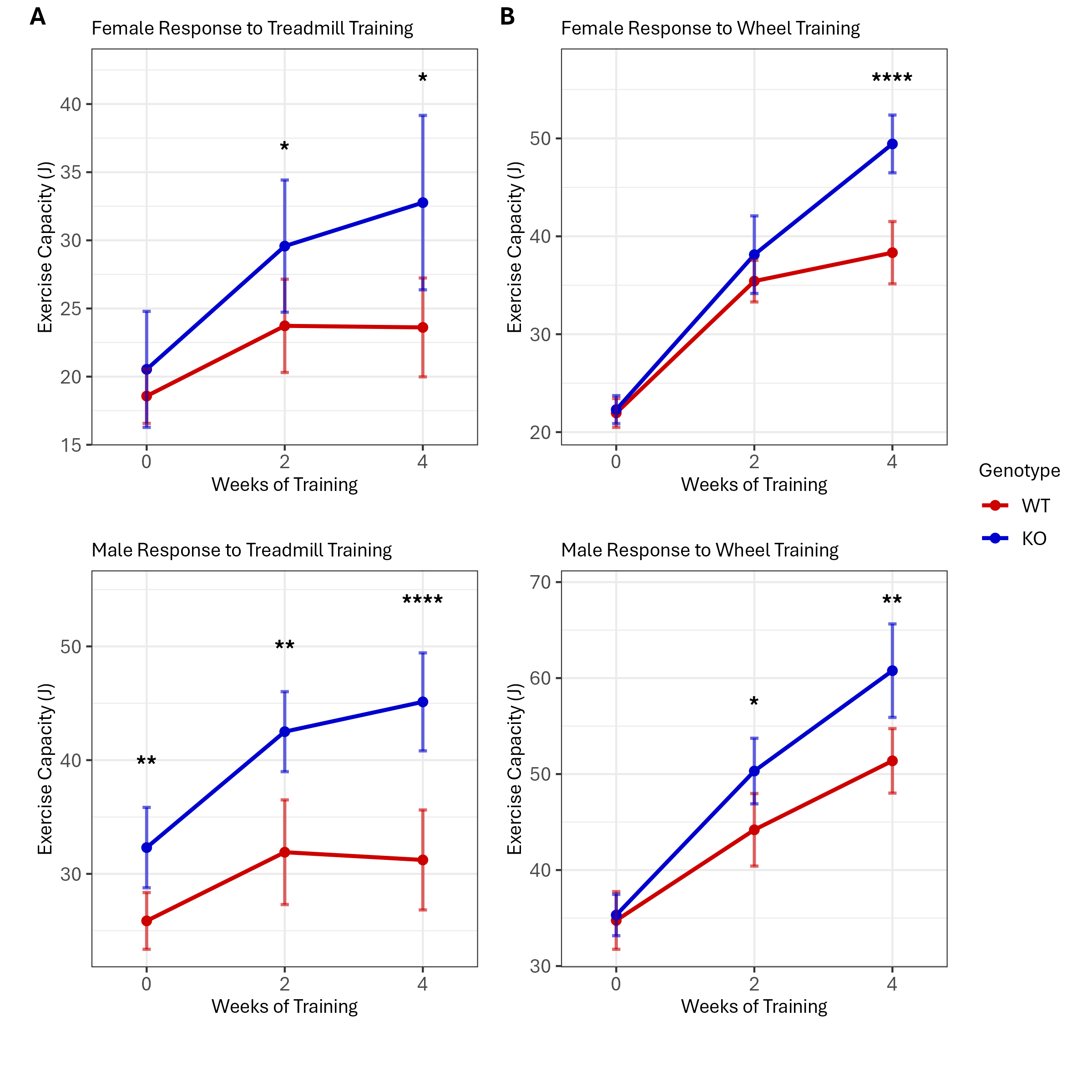

**Figure S4. Causal impact of IGFBP7 knockout on trainability.** Plots depict absolute exercise capacity as a function of training time. (**A**) The effect of matched treadmill training on exercise capacity, stratified by sex; N=8 to 14 animals per group. (**B**) The effect of voluntary wheel training on exercise capacity, stratified by sex; N=7 to 11 animals per group. Error bars indicate 95% confidence intervals. ****, P<0.0001; **, P<0.01; *, P<0.05. WT indicates wild-type; KO, IGFBP7 knockout.

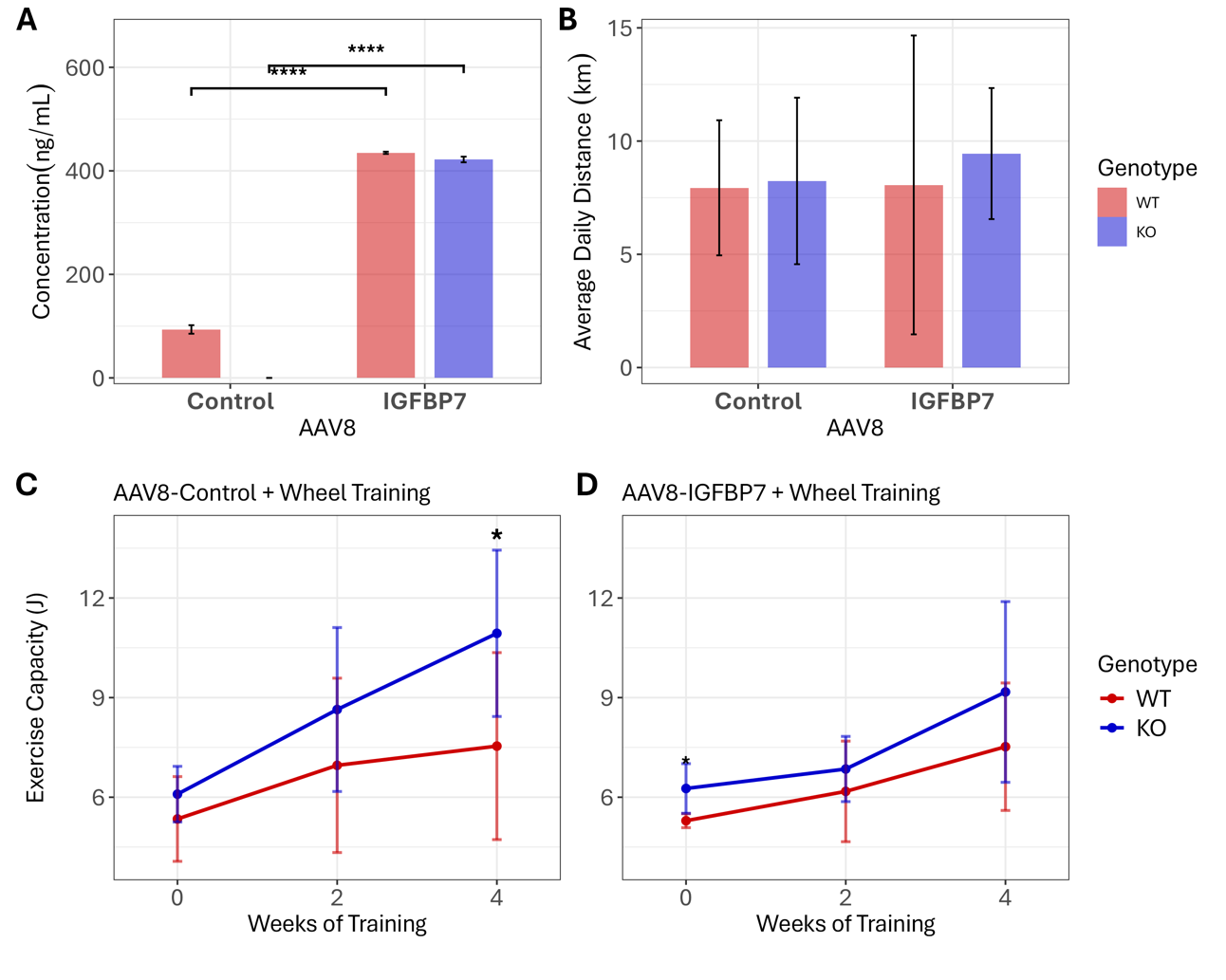

**Figure S5. Effect of IGFBP7 overexpression on trainability. (A)** Circulating IGFBP7 levels resulting from AAV8-IGFBP7 overexpression vs an AAV8-Control vector, in IGFBP7 KO and WT female mice. **(B)** Average daily running distance by genotype and AAV8 exposure. (**C-D)** Effect of AAV8 mediated overexpression of IGFBP7 on fitness change from wheel running in female mice (N=4-5 per group). A significant difference in 4-week exercise capacity was seen between genotypes in AAV8-Control treated mice, P=0.03 (t-test), but not in AAV8-IGFBP7 overexpressing mice. A significant difference in the slope of the training effect was seen between genotypes in AAV8-Control treated mice, 1.2 J/week in KO vs 0.55 J/week in WT, P=0.007 (linear mixed model); no significant difference in slopes was seen in AAV8-IGFBP7 overexpressing mice. Error bars indicate 95% confidence intervals. * P < 0.05, **** P < 0.001. AAV8 indicates adeno-associated virus serotype 8.

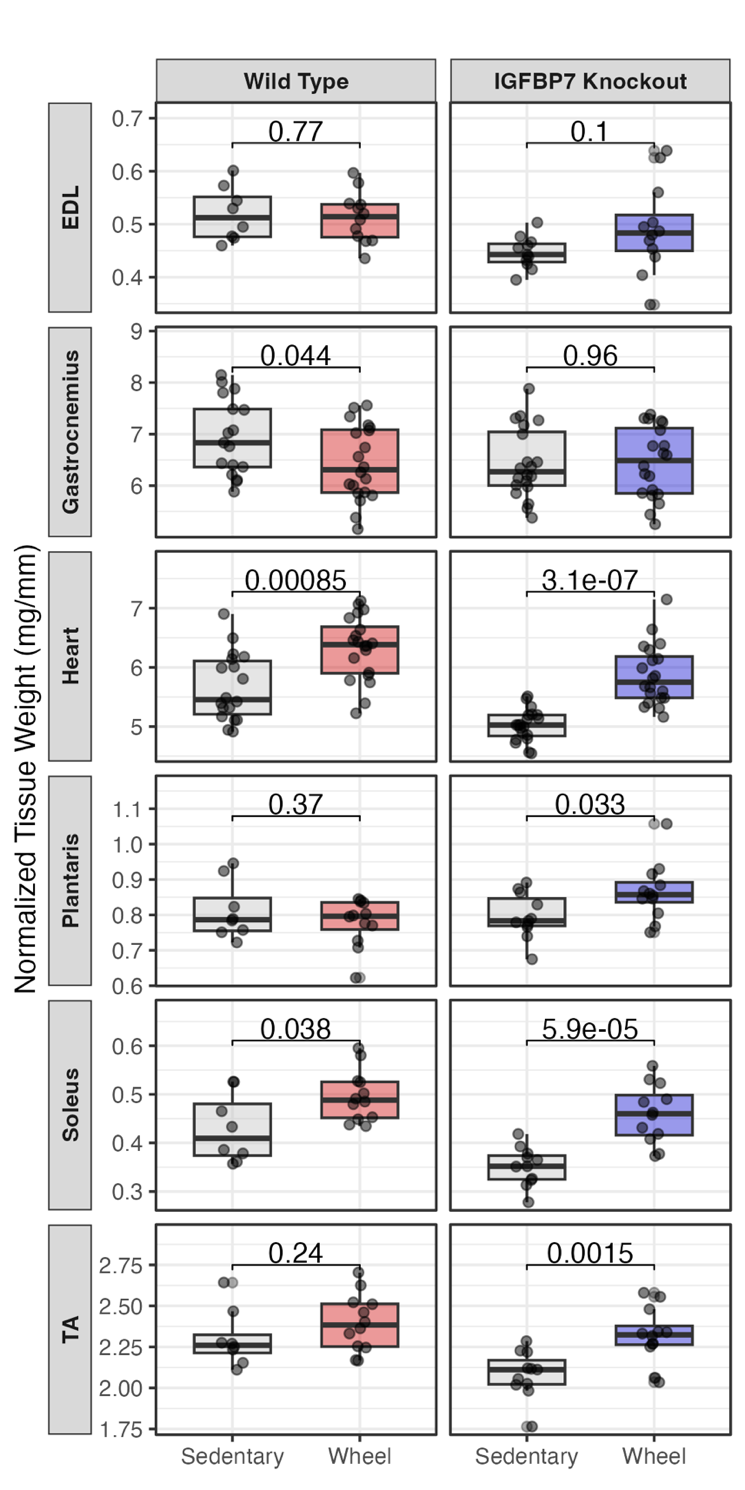

**Figure S6. Muscle weights in sedentary and trained female mice.** Comparison of muscle weights between sedentary and wheel-trained female mice, stratified by genotype. Muscle weight was normalized to tibia length. The P-values for differences in muscle weight as assessed by t-test are annotated on the cross bars. Error bars indicate 95% confidence intervals. EDL indicates extensor digitorum longus; TA, tibialis anterior.

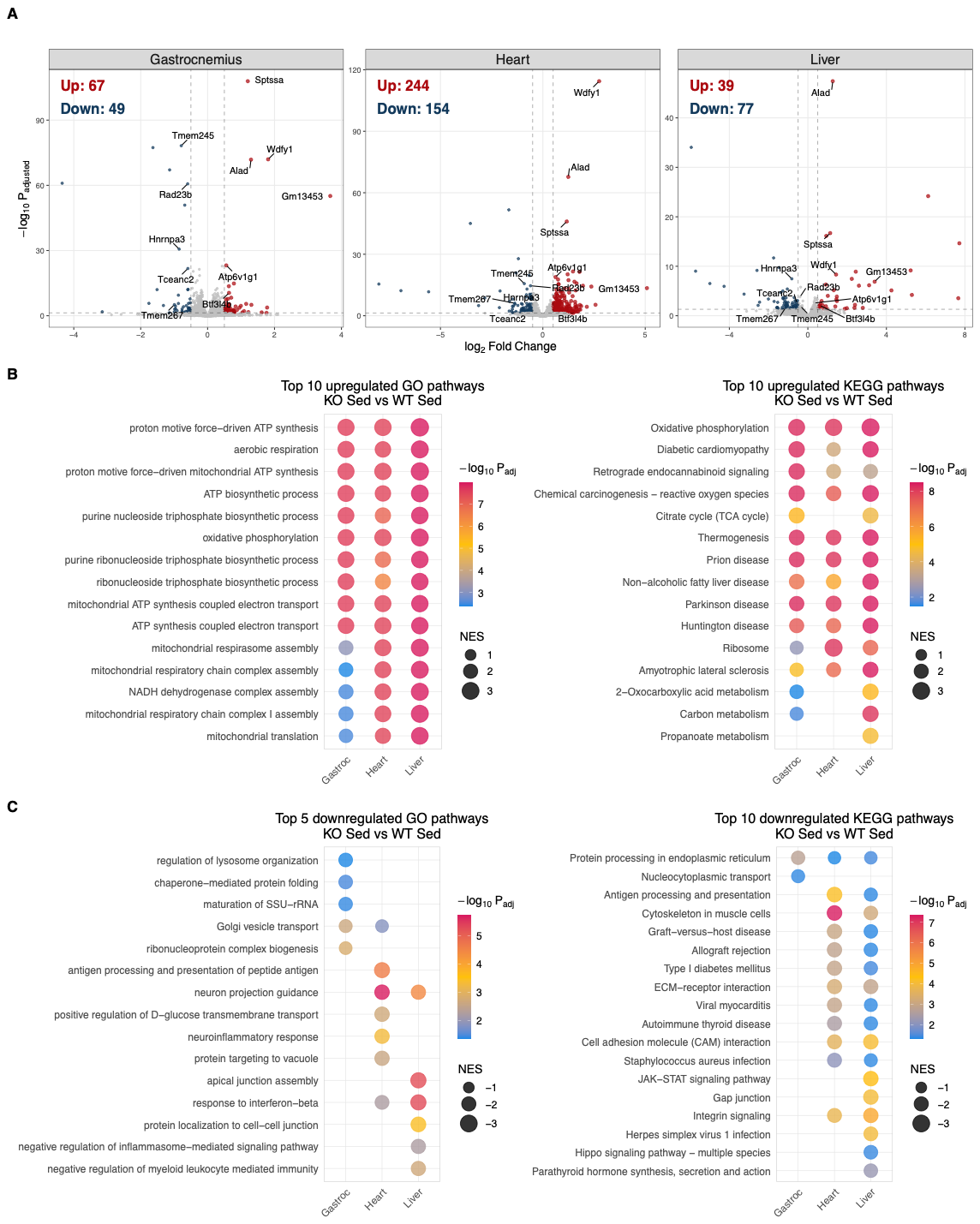

**Figure S7. Differential expression analysis between KO and WT sedentary groups across tissues.** (**A**) Volcano plots showing the DEGs between KO sedentary and WT sedentary groups across gastrocnemius, heart and liver from female mice. Common DEGs are annotated. (**B**) Top positively enriched pathways in each tissue. (**C**) Top negatively enriched pathways. Both GO biological processes and KEGG pathways were analyzed by Gene Set Enrichment Analysis (FDR< 0.05). KO indicates knockout; WT, wild-type; Sed, sedentary; DEG, differentially expressed gene; FC, fold-change; FDR, false-discovery rate; NES, normalized enrichment score; GO, Gene Ontology; KEGG, Kyoto encyclopedia of genes and genomes.

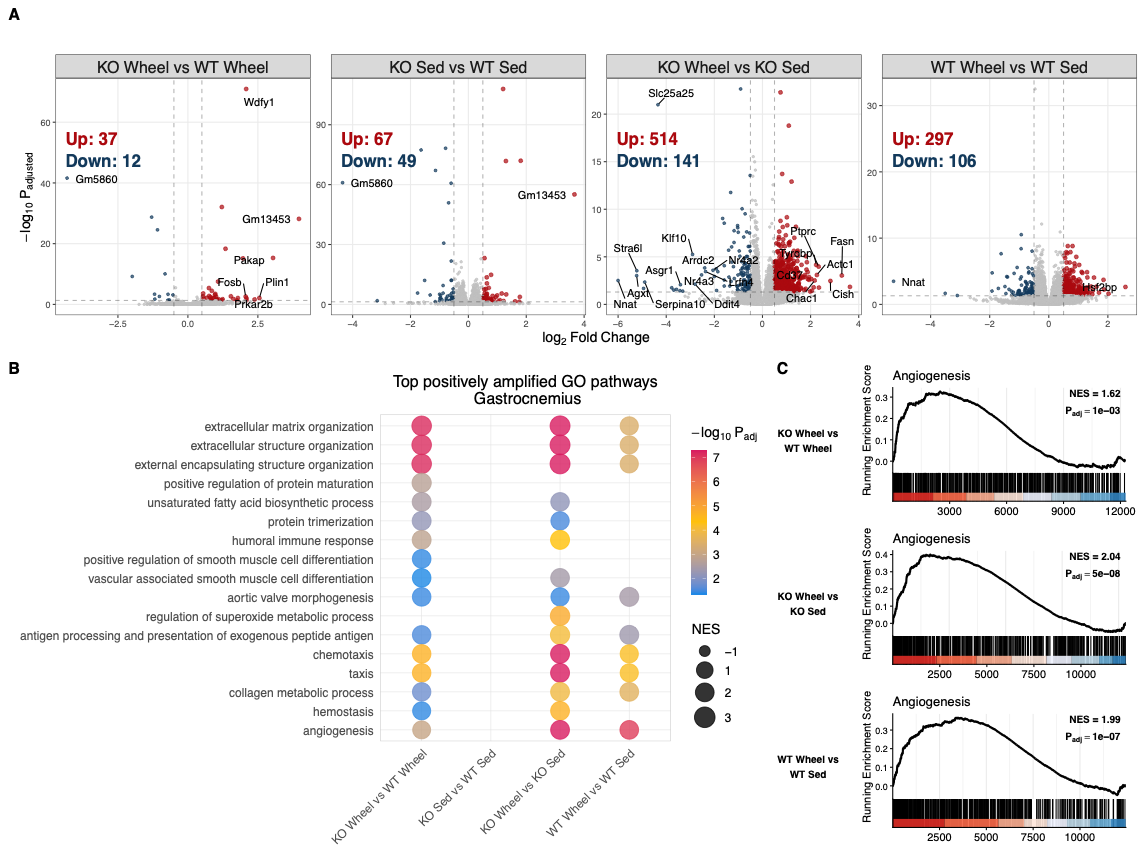

**Figure S8. Transcriptome analysis of amplified trainability of IGFBP7 KO in gastrocnemius.** (**A**) Volcano plots showing DEGs across genotype and training comparisons in gastrocnemius from female mice (|log_2_FC| > 0.5, FDR < 0.05). (**B**) Amplified pathway comparisons. Top 10 pathways most enriched at peak training (KO Wheel vs WT Wheel) and top 10 pathways enriched by training in KO mice (KO Wheel vs KO Sedentary). Only significantly enriched pathways are plotted for each of the 4 comparisons (FDR<0.05). (**C**) GSEA plot for angiogenesis pathway for three comparisons. KO indicates knockout; WT, wild-type; Sed, sedentary; DEG, differentially expressed gene; FC, fold-change; FDR, false-discovery rate; NES, normalized enrichment score; GO, Gene Ontology; GSEA indicates genes set enrichment analysis.

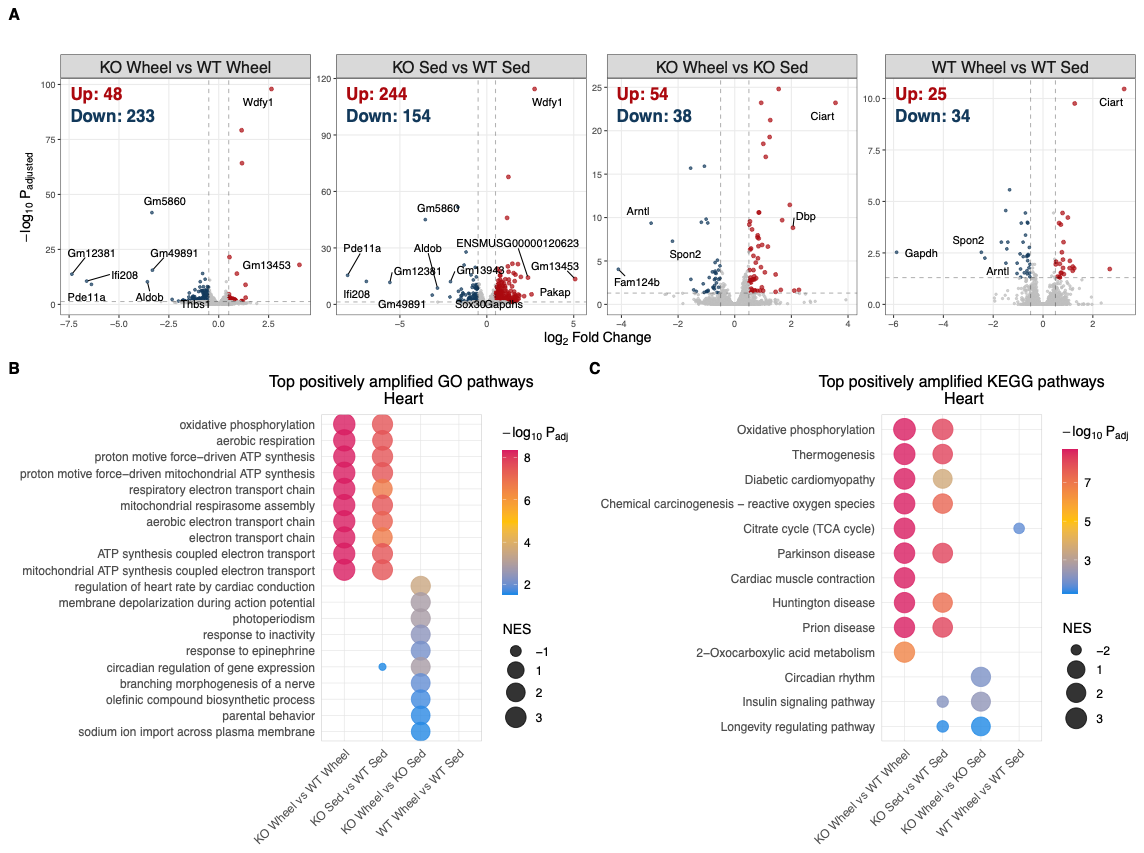

**Figure S9. Cardiac transcriptional programs associated with amplified trainability in IGFBP7 KO mice.** (**A**) Volcano plot showing DEGs across genotype and training comparisons in heart from female mice (|log_2_FC| > 0.5, FDR < 0.05). (**B**) Comparisons to infer amplification of GO-biological process pathways. Top 10 pathways most enriched at peak training (KO Wheel vs WT Wheel) and top 10 pathways enriched by training in KO mice (KO Wheel vs KO Sedentary). Only pathways significantly enriched in a given comparison are displayed in the dot plot (FDR<0.05). (**C**) Identical to (B), but for KEGG pathways. KO indicates knockout; WT, wild-type; Sed, sedentary; DEG, differentially expressed gene; FC, fold-change; FDR, false-discovery rate; NES, normalized enrichment score; GO, Gene Ontology; KEGG, Kyoto encyclopedia of genes and genomes.

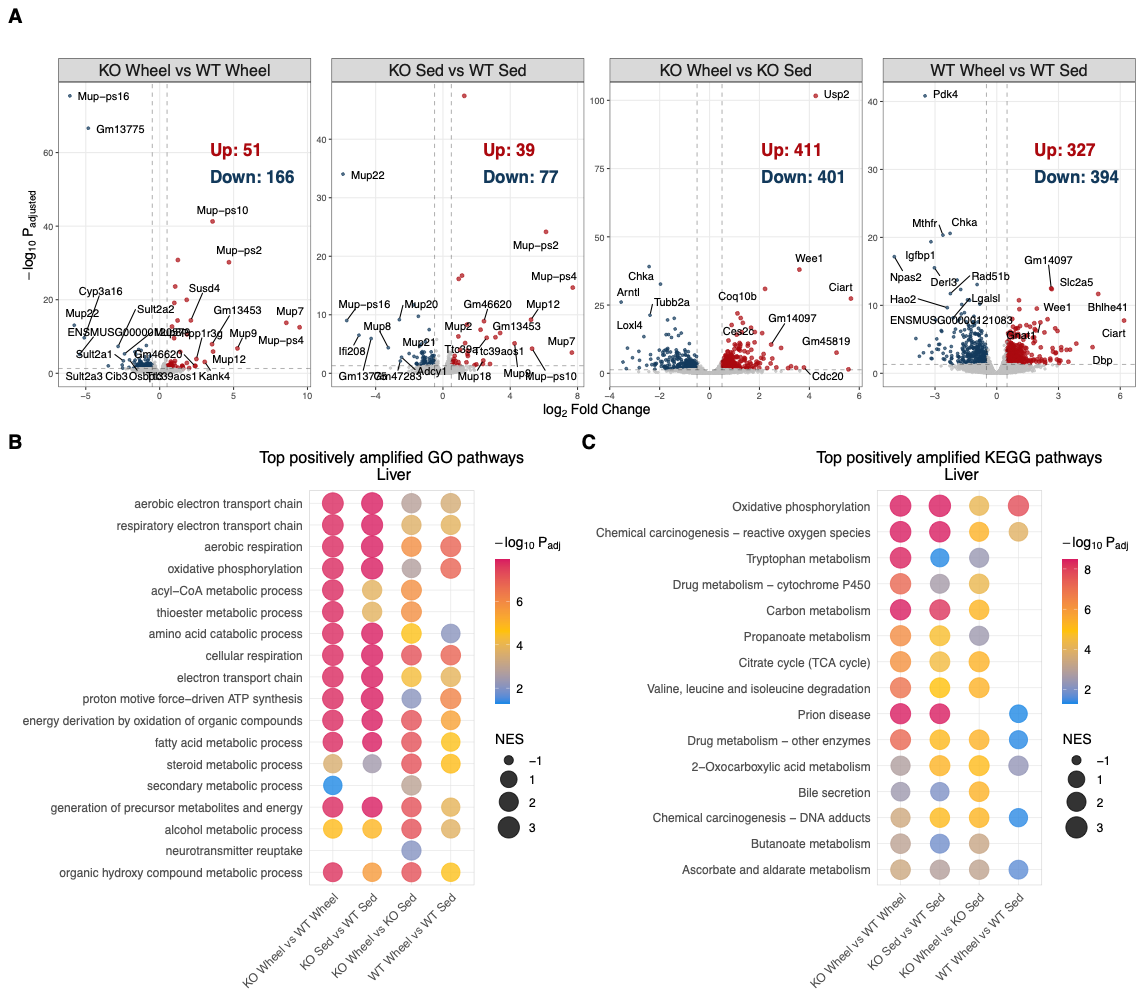

**Figure S10. Hepatic transcriptional programs associated with amplified trainability in IGFBP7 KO mice.** (**A**) Volcano plot showing DEGs across genotype and training comparisons in liver from female mice (|log_2_FC| > 0.5, FDR < 0.05). (**B**) Comparisons to infer amplification of GO-biological process pathways. Top 10 pathways most enriched at peak training (KO Wheel vs WT Wheel) and top 10 pathways enriched by training in KO mice (KO Wheel vs KO Sedentary). Only pathways significantly enriched in a given comparison are displayed in the dot plot (FDR<0.05). (**C**) Identical to (B), but for KEGG pathways. KO indicates knockout; WT, wild-type; Sed, sedentary; DEG, differentially expressed gene; FC, fold-change; FDR, false-discovery rate; NES, normalized enrichment score; GO, Gene Ontology; KEGG, Kyoto encyclopedia of genes and genomes.

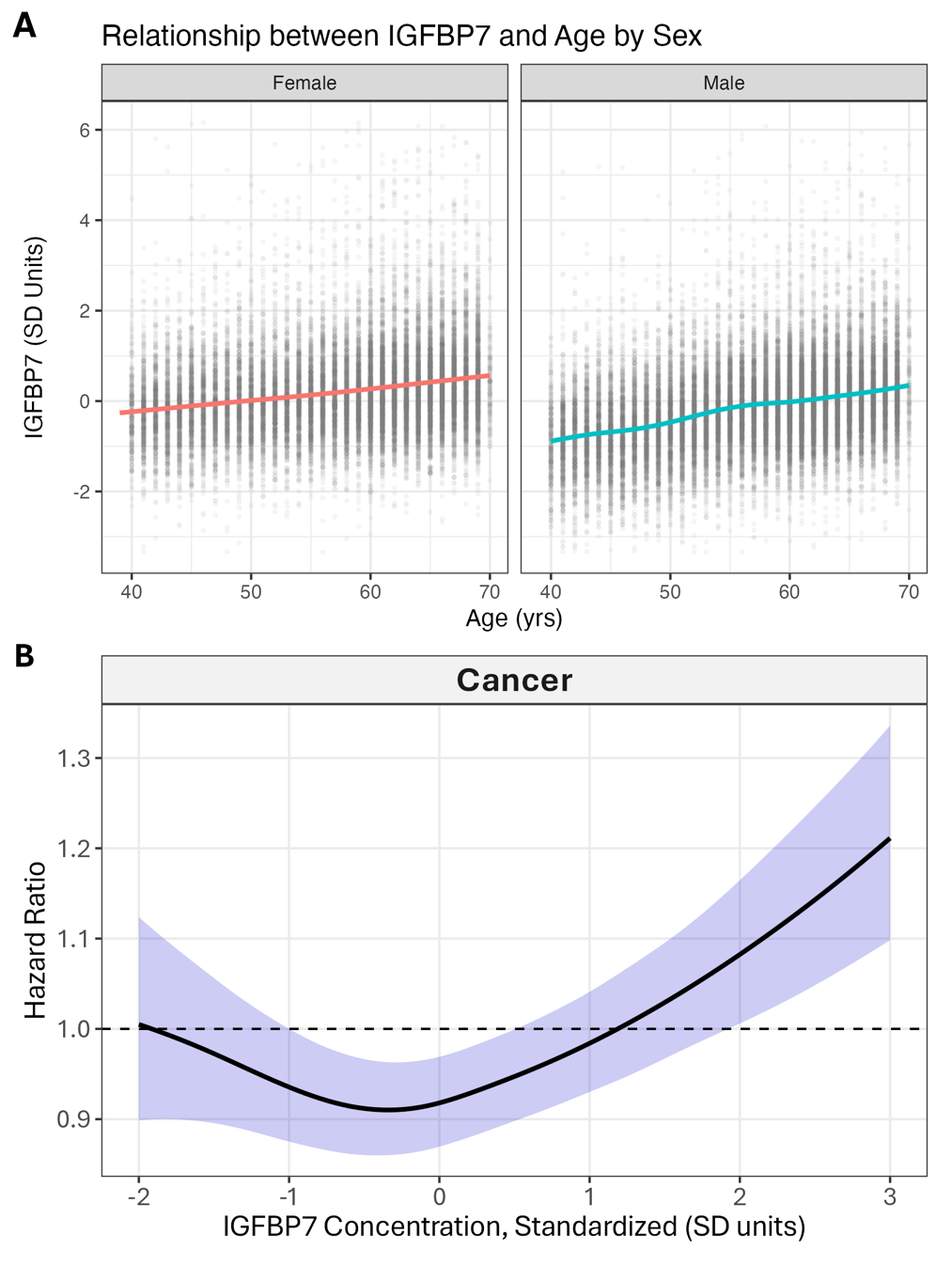

**Figure S11. (A) Age and IGFBP7 levels in the UK Biobank.** Relationship between IGFBP7 level and age in the UK Biobank. IGFBP7 was measured by the OLink platform. **(B) Cancer incidence and IGFBP7 levels.** Relationship between IGFBP7 level and incident cancer in a proportional hazards model adjusted for age and sex, fitted to participant data from the UK Biobank. IGFBP7 was measured by the OLink platform. Error bands indicate 95% confidence bands.

**Supplementary Data Descriptions**

**Data S1: Proteomics**

Tables of plasma proteins found to have differential abundance across comparisons such as High vs Low responders, baseline vs post-training, and pre- vs post-acute exercise. A table with the proteins whose baseline level predicts VO2 response in the discovery cohort is also included.

**Data S2: Differentially Expressed Genes**

Tables of differentially expressed genes (DEGs) across comparisons such as IGFBP7 knockout vs wild-type stratified by training status, sedentary vs trained stratified by genotype. Tables of DEGs across each of three tissues – gastrocnemius, heart, and liver – are included.

**Data S3: Pathway Enrichment**

Tables of enriched pathways corresponding to the comparisons in Data S2. Pathways are reported from both the Gene Ontology (GO) database and Kyoto encyclopedia of genes and genomes (KEGG).

**Data S4: Pathway Amplification**

Tables of pathways whose pattern of enrichment can be classified as differentially amplified by training in IGFBP7 KO mice, either positively or negatively. Both GO and KEGG pathway amplification are included, across gastrocnemius, heart, and liver. A pathway can be classified as amplified by each of four criteria, referred to as amplification types (Methods) and reported in a table column. When pathway amplification is supported by two or more amplification criteria, the numerical statistics reported in the pathway row correspond to the amplification type with the maximum absolute normalized enrichment score.
